# Rigorous benchmarking of T cell receptor repertoire profiling methods for cancer RNA sequencing

**DOI:** 10.1101/2022.03.31.22273249

**Authors:** Kerui Peng, Theodore Scott Nowicki, Katie Campbell, Dandan Peng, Anish Nagareddy, Yu-Ning Huang, Aaron Karlsberg, Zachary Miller, Jaqueline Brito, Victoria M. Pak, Malak S. Abedalthagafi, Amanda M. Burkhardt, Houda Alachkar, Antoni Ribas, Serghei Mangul

**Affiliations:** Department of Clinical Pharmacy, School of Pharmacy, University of Southern California, Los Angeles, CA, USA; Department of Pediatrics, Division of Pediatric Hematology/Oncology, University of California, Los Angeles, CA, USA; Department of Microbiology, Immunology, & Molecular Genetics, University of California, Los Angeles, CA, USA; Department of Medicine, Division of Hematology-Oncology, University of California, Los Angeles, CA, USA; Department of Quantitative and Computational Biology, Dana and David Dornsife College of Letters, Arts and Sciences, University of Southern California, Los Angeles, CA, USA; Viterbi School of Engineering, University of Southern California, Los Angeles, CA, USA; Department of Pharmaceutical Sciences, School of Pharmacy, University of Southern California, Los Angeles, CA, USA; Emory Nell Hodgson School of Nursing, Emory University, Atlanta, GA, USA; Department of Epidemiology, Rollins School of Public Health, Emory University, Atlanta, GA, USA; Genomics Research Department, Saudi Human Genome Project, King Fahad Medical City and King Abdulaziz City for Science and Technology, Riyadh, Saudi Arabia; King Salman Center for Disability Research, Riyadh 12512, Saudi Arabia; College of Medicine, Imam Mohammad Ibn Saud Islamic University (IMSIU), Riyadh, Saudi Arabia; Departments of Medicine (Hematology-Oncology), Surgery (Surgical Oncology) and Molecular & Medical Pharmacology, University of California, Los Angeles, CA, USA; Department of Clinical Pharmacy, School of Pharmacy, University of Southern California, 1540 Alcazar Street, Los Angeles, CA 90033, USA

## Abstract

The ability to identify and track T cell receptor (TCR) sequences from patient samples is becoming central to the field of cancer research and immunotherapy. Tracking genetically engineered T cells expressing TCRs that target specific tumor antigens is important to determine the persistence of these cells and quantify tumor responses. The available high-throughput method to profile T cell receptor repertoires is generally referred to as TCR sequencing (TCR-Seq). However, the available TCR-Seq data is limited compared to RNA sequencing (RNA-Seq). In this paper, we have benchmarked the ability of RNA-Seq-based methods to profile TCR repertoires by examining 19 bulk RNA-Seq samples across four cancer cohorts including both T cell rich and poor tissue types. We have performed a comprehensive evaluation of the existing RNA-Seq-based repertoire profiling methods using targeted TCR-Seq as the gold standard. We also highlighted scenarios under which the RNA-Seq approach is suitable and can provide comparable accuracy to the TCR-Seq approach. Our results show that RNA-Seq-based methods are able to effectively capture the clonotypes and estimate the diversity of TCR repertoires, as well as to provide relative frequencies of clonotypes in T cell rich tissues and monoclonal repertoires. However, RNA-Seq-based TCR profiling methods have limited power in T cell poor tissues, especially in polyclonal repertoires of T cell poor tissues. The results of our benchmarking provide an additional appealing argument to incorporate the RNA-Seq into immune repertoire screening of cancer patients as it offers broader knowledge into the transcriptomic changes that exceed the limited information provided by TCR-Seq.

## Introduction

Immunotherapy is an effective approach to treat a variety of advanced malignancies^1,2^. The success of immunotherapy relies in part on the presence of CD8+ cytotoxic T cells, which recognize tumor antigens via their T cell receptors (TCRs), and then induce targeted cell apoptosis^1^. The ability to characterize the TCR repertoire in patient samples is increasingly central to the field of immunotherapy and cancer research^3,4^. Targeted TCR Sequencing (TCR-Seq) approaches are currently used to measure the diversity and clonality of the TCR repertoire^5–9^, which can be affected by immune checkpoint inhibitors and can serve as a surrogate measure of effectiveness and overall prognosis^10–12^. Furthermore, the ability to track genetically engineered T cells expressing chimeric TCRs that target specific tumor antigens is important to determine the persistence of these cells and corresponding clinical responses. However, TCR-Seq is not frequently done compared to the RNA-Seq, thus there is a significant amount of RNA-Seq data available that can be used to extract TCR data. RNA-Seq-based TCR profiling methods have been developed to bridge the gap^13–15^. For example, a recent study demonstrated that MiXCR could detect all TCRβ sequences with relative frequencies greater than 0.15% in one of T cell rich tissues^16^. However, despite the great promise of RNA-Seq-based TCR profiling methods, such methods were not systematically benchmarked and they were validated in extremely small numbers of samples and limited scenarios. Thus, the biomedical communities remain uninformed regarding the advantages and limitations of these RNA-Seq-based TCR profiling methods. Additionally, the feasibility of applying these methods across various cancer tissue types and TCR repertoire types remains unknown.

Here we performed a comprehensive benchmarking of existing RNA-Seq-based TCR profiling methods. The performance of these methods was investigated by using TCR-Seq as a gold standard across T cell rich and poor tissues from different cancer tissue types and immune repertoire types. We have carefully examined the scenarios of monoclonal (repertoires are dominated by one or a few clonotypes with high frequencies) and polyclonal (repertoires are composed of clonotypes with frequencies nearly evenly distributed) TCR repertoires under which such tools can leverage RNA-Seq data and provide reliable estimates of TCR repertoires, which is currently unknown. Our results show that RNA-Seq-based TCR profiling methods are able to effectively capture the majority of TCR-Seq confirmed clonotypes in monoclonal repertoires. These methods are also capable of precisely estimating the overall TCR repertoire diversity and clonotype frequencies in T cell rich monoclonal repertoires. However, in the case of monoclonal repertoires, the small number of receptor reads that capture major clonotypes is enough to estimate diversity even in T cell poor tissues. Moreover, bulk RNA-Seq stored both TCRα and TCRβ chain information, these methods offered the possibility to characterize both TCRα and TCRβ repertoires. However, cautions need to be taken for T cell poor tissues as the results typically were less accurate because of the lack of ability of RNA-Seq-based methods to detect clonotypes with low frequencies. To conclude, we have examined the ability of RNA-Seq-based TCR profiling methods in different types of tissues and repertoires, thus providing a comprehensive guide in which tissues RNA-Seq-based TCR profiling methods are feasible to deliver comparable results to targeted TCR-Seq.

## Results

### Gold standard datasets

We assembled the largest multi-cohort dataset which was composed of 19 samples and 5 tissue types (Table 1). Peripheral blood mononuclear cells (PBMC) (N=5) and melanoma biopsy samples (N=9) were generated from patients at UCLA enrolled in transgenic NY-ESO-1 TCR adoptive cell therapy (IRB#12-000153 and 13-001624)^17^ and PD-1 blockade clinical trials (IRB#11-003066)^12^, as previously described. The studies were conducted in accordance with local regulations, the guidelines for Good Clinical Practice, and the principles of the Declaration of Helsinki. Renal clear cell carcinoma (N=3)^18^ samples were acquired from The Cancer Genome Atlas (TCGA), melanoma specimens from the ileocecal lymph node (N=1) and the small intestine (N=1)^16^ were acquired from the Sequence Read Archive (SRA). For the PBMC samples, three were from patients that were transduced with a retroviral vector containing the NY-ESO-1 TCR^17^, while another two were from patients being treated for melanoma^12^. All the samples were sequenced by both TCR-Seq (only for TCRβ chain) and RNA-Seq. The average number of RNA-Seq reads ranged from 66 million to 88.3 million. The average numbers of TCR-Seq reads ranged from 0.01 million to 3.26 million reads (Table 1, Supplementary Table 1, Supplementary Fig.1a-b). The number of clonotypes detected by TCR-Seq varied across different tissue types. Kidney samples had the lowest average number of clonotypes (1,917) in the tissue samples analyzed. Note that the lymph node sample has an extremely high number of TCRβ clonotypes (202,869) compared to all other samples (Supplementary Fig.1c). Importantly, the cohorts we used correspond to various tissue types with substantially different levels of T cells and repertoire types. Samples with high levels of T cells were from PBMC and lymph nodes, which were considered as T cell rich tissues. While low levels of T cells were the samples from the renal cells, small intestine, and melanoma biopsies, which were considered as T cell poor tissues.

**Table 1.** Overview of the gold standard dataset. We had different cohorts from five different tissue types in total. We documented the average number of TCR-Seq reads in million (indicated in the column “Average number of TCR-Seq reads”), the average number of RNA-Seq reads in million (indicated in the column “Average number of RNA-Seq reads”), RNA-Seq read length (indicated in the column “Length of RNA-Seq reads”), the fraction of T cells in certain tissue type (indicated in the column “Fraction of T cells in tissue”), the number of samples from each tissue type (indicated in the column “Number of samples “), the number of monoclonal samples and polyclonal samples from each tissue type (indicated in the column “Number of monoclonal/polyclonal samples”).

| Tissue type | Number of TCR-Seq reads (millions) (mean±SD) | Number of RNA-Seq reads (millions) (mean±SD) | Length of RNA-Seq reads | Fraction of T cells in tissue | Number of samples | Number of monoclonal / polyclonal samples | Resources |
| --- | --- | --- | --- | --- | --- | --- | --- |
| PBMC | 0.122±0.1 | 66.8±24.4 | 150bp | High | 5 | 3/2 | In-house |
| Melanoma biopsies | 0.951±0.523 | 74.7±15.9 | 100bp | Low | 9 | 1/8 | In-house |
| Renal cell | 0.0102±0.007 | 88.3±28 | 50bp | Low | 3 | 0/3 | TCGA |
| Lymph node | 3.26 | 66 | 100bp | High | 1 | 0/1 | SRA |
| Small intestine | 3.05 | 72.7 | 100bp | Low | 1 | 0/1 | SRA |

We used Shannon Diversity Index (SDI) to classify TCR repertoires as monoclonal or polyclonal. The minimal value of SDI is 0, which value closer to 0 corresponds to the increased monoclonality of the TCR population. We defined the samples with SDI less than 2 as monoclonal samples, otherwise polyclonal samples (Supplementary Fig.2). The three PBMC samples that were transduced with a retroviral vector containing the NY-ESO-1 TCR^17^ and one melanoma biopsy sample were monoclonal samples, while all other samples were polyclonal. In our dataset, the clonotypes with frequencies greater than 0.01 (hyperexpanded clonotypes), which were usually only one or a few TCRβ clonotypes, on average composed 91.08% of all TCRβ clonotypes in monoclonal samples, while the hyperexpanded clonotypes only composed 24.36% of all TCRβ clonotypes in polyclonal samples (Supplementary Fig.3).

### Choice of RNA-Seq-based TCR profiling methods

All the available RNA-Seq-based repertoire profiling methods that are designed to assemble the complementarity determining regions 3 (CDR3) from RNA-Seq data were included in this benchmarking study, which includes MiXCR^13^, ImReP^14^, and TRUST4^15^. Notably, MiXCR supports various sequencing technologies, while ImReP and TRUST4 are designed mainly for RNA-seq data (Table 2 and Table 3). The input format varied across tools, MiXCR requires RNA-Seq raw reads, ImReP accepts only aligned reads, TRUST4 accepts both raw reads and aligned reads. The details on how the RNA-Seq-based methods were run are available in Supplementary Table 2.

**Table 2.** Overview of evaluated RNA-Seq-based TCR profiling methods’ parameters, publication details, and technical characteristics. RNA-Seq-based TCR profiling methods are sorted by the year of publication (indicated in the column “Published year”). We documented the name of the methods (indicated in the column “Method”), the version of methods that were used in this benchmarking project (indicated in the column “Version”), the alignment algorithms (indicated in the column “Alignment algorithm”), the programming language (indicated in the column “Programming language”), the input file type (indicated in the column “Input file type”), the types of sequencing data accepted by the method (indicated in the column “Types of sequencing data”), the organism accepted by the method (indicated in the column “Organism”), the journal that the method was published in (indicated in the column “Journal”), the computational tools that the method was compared to in the publication (indicated in the column “In the publication compared to”), the method webpage (indicated in the column “Method webpage”).

| Method | Version | Alignment algorithm | Published year | Programming language | Input file type | Types of supported data | Organism | Journal | In the publication compared to | Webpage |
| --- | --- | --- | --- | --- | --- | --- | --- | --- | --- | --- |
| MiXCR | 3.0.13 | modified Smith-Waterman and Needleman-Wunsch | 2015 | Java | FASTA, FASTQ, FASTQ [.gz] Paired-end FASTQ [.gz] | AIRR-seq, RNA-seq | Human, mouse, rat | Nature Methods | MiTCR, Decombinator, IgBLAST, IMGT/HighV QUEST | https://milaboratory.com/software/mixcr/ |
| ImReP | 1.0 | Not required | 2020 | Python | BAM | RNA-seq | Human, mouse | Nature Communications | MiXCR, IMSEQ, IgBLAST | https://github.com/Mangul-Lab-USC/ImReP |
| TRUST4 | 1.0.2 | Seed-extension | 2021 | C/C++ | BAM, FASTQ | RNA-seq | Human | Nature Methods | MiXCR, CATT, TRUST3 | https://github.com/liulab-dfci/TRUST4 |

### RNA-Seq-based TCR profiling methods were able to successfully capture TCRβ monoclonal repertoires across various T cell rich tissues

First, we have investigated the ability of RNA-Seq-based TCR profiling methods to characterize TCRβ repertoires across different repertoire types and tissue types. The capturing ability of the RNA-Seq-based methods was examined based on the sum of the TCR-Seq confirmed TCRβ clonotypes frequencies that each method was able to capture. Each distinct amino acid sequence was considered as one unique clonotype in the TCRβ repertoire.

The surveyed RNA-Seq-based methods were able to capture 93.18% of clonotypes in T cell rich monoclonal samples and 76.61% in T cell poor monoclonal samples on average (Fig.1a-c). Similar to the previous study^16^, we observed that MiXCR and TRUST4 were able to capture the entire repertoire of T cell rich monoclonal samples composed from clonotypes with frequency as low as 0.03%. All surveyed methods captured the repertoire composed of clonotypes with frequencies above 0.036% in T cell rich monoclonal samples and clonotypes with frequencies above 2.66% in T cell poor monoclonal samples (Fig.1a-c). They were able to capture the entire repertoire composed of clonotypes with frequencies above 1.553% in T cell rich polyclonal samples (Fig.1d-f). However, only TCRβ clonotypes with relatively high frequencies in the T cell poor polyclonal samples were captured by ImRep and TRUST4, but not by MiXCR. Among ImRep and TRUST4, TRUST4 achieved the best performance compared to ImReP in the T cell poor polyclonal repertoires, with TRUST4 being able to capture all the TCRβ clonotypes with frequencies greater than 10.1%, while this threshold was 40.5% for ImReP (Fig.1d-f). One reason that may have caused this was that on average TCRβ derived reads from RNA-Seq reads in T cell poor tissues were much smaller than those in T cell rich tissues (4.1 vs. 655.3 TCR derived reads per million RNA-Seq reads) (Supplementary Table 3, Supplementary Table 4, Supplementary Fig.4). The results for individual samples were shown in Supplementary Fig.5.

**Figure 1.**
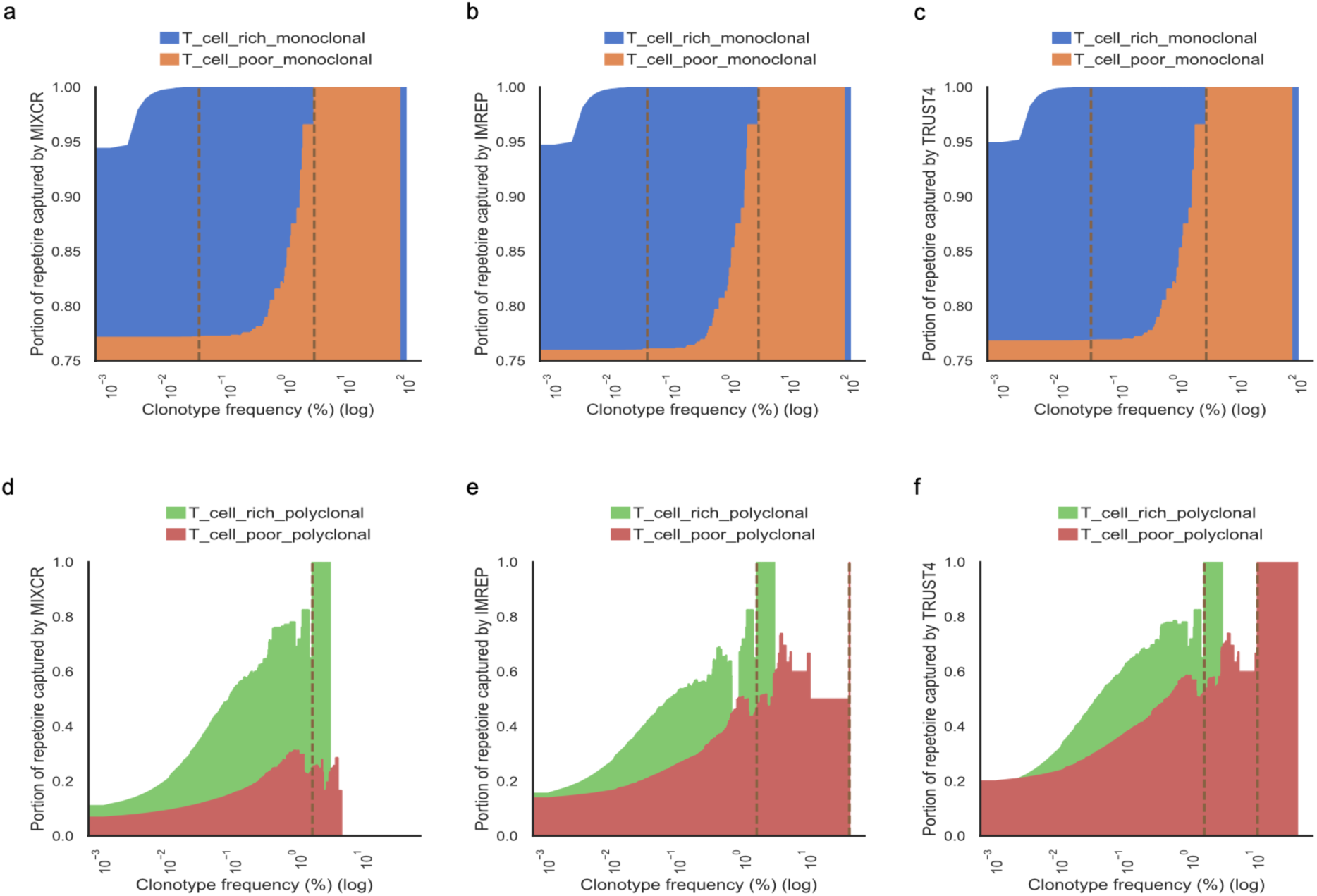
The ability to capture TCR-Seq confirmed clonotypes by RNA-Seq-based methods in monoclonal samples (a-c) and polyclonal samples (d-f). The x-axis corresponds to TCR-seq confirmed clonotypes with a frequency of Z on a log scale. The y-axis corresponds to the fraction of assembled TCR repertoire by RNA-Seq-based methods with clonotype frequency greater than Z. The brown dash lines indicate the minimal clonotype frequency above which the entire repertoire is captured. Area plots show the proportion of the total TCR repertoire captured by MiXCR, ImReP, and TRUST4. Results from T cell rich monoclonal samples, T cell poor monoclonal samples, T cell rich polyclonal samples, and T cell poor polyclonal samples are shown in blue, orange, green, and red, respectively.

We further examined the ability of the surveyed methods to capture the most abundant clonotypes (top five and top ten clonotypes in each sample) in T cell rich tissues. MiXCR and TRUST4 were able to capture the entire set of the top five clonotypes in T cell rich monoclonal samples while ImReP was able to capture 93.3% of those clonotypes. For the top ten clonotypes in T cell rich monoclonal samples, MiXCR, ImReP, and TRUST4 were able to capture 93.3%, 86.7%, and 96.7%, respectively. While in the T cell rich polyclonal samples, 66.7% of the top five clonotypes and 52.2% of the top ten clonotypes were captured by these methods on average.

### RNA-Seq based TCR profiling methods were able to effectively estimate diversity in T cell rich tissues

We evaluated the ability to effectively estimate the diversity of TCR repertoire by RNA-Seq-based methods. The diversity is estimated by Shannon Diversity Index. All three RNA-Seq-based methods provided reliable estimates of diversity compared with the diversity estimates from TCR-Seq-based results in T cell rich samples (MiXCR: r=0.95, *p*=0.003; ImReP: r=0.9, *p*=0.016; TRUST4: r=0.94, *p*=0.0055) (Fig.2a-c).

**Figure 2.**
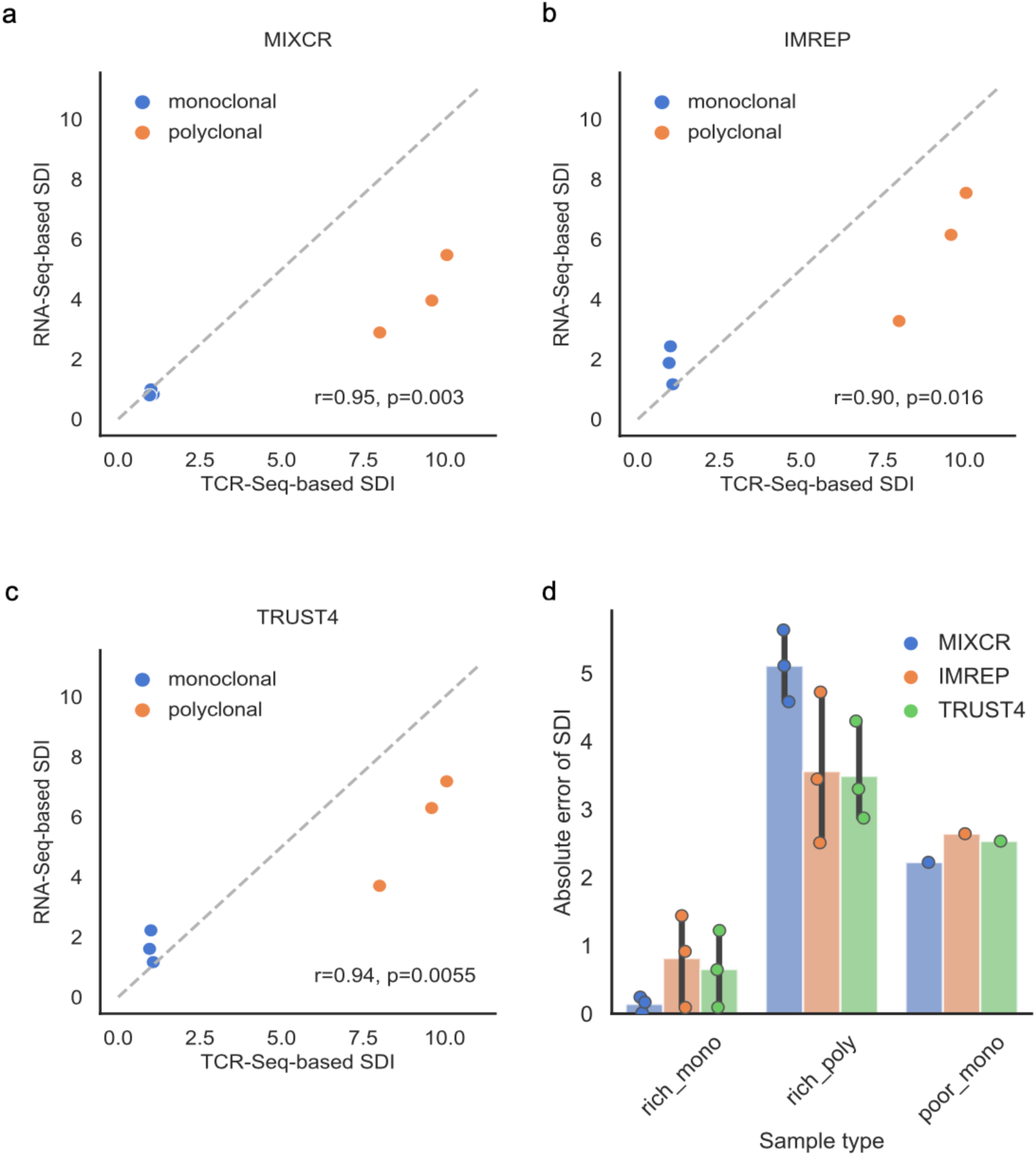
TCR-Seq-based TCRβ diversity and its comparison with RNA-Seq-based TCRβ diversity. a-c. Correlation of diversity estimated based on RNA-Seq data (y-axis) and TCR-Seq data (x-axis) by Shannon Diversity Index (SDI) in T cell rich tissues. Blue represents monoclonal samples, orange represents polyclonal samples. Pearson correlation coefficients and the corresponding p-values are calculated and reported. d. Bar plot and scatter plot of absolute error between diversity estimated based on RNA-Seq data and TCR-Seq data in T cell rich monoclonal samples (rich_mono), T cell rich polyclonal samples (rich_poly), and T cell poor monoclonal samples (poor_mono). The results from MiXCR are represented in blue, the results from ImReP are represented in orange, the results from TRUST4 are represented in green.

To further compare the ability of methods to estimate diversity across various types of tissues and repertoire types, we also used absolute error to examine the feasibility of the methods to estimate diversity. The differences between the Shannon Diversity Index estimated from TCR-Seq results and RNA-Seq TCR derived results were measured using the absolute error. The average absolute error of diversity estimation from RNA-Seq-based methods compared to TCR-Seq results in T cell rich tissues was 2.29. The best performance was achieved by TRUST4 with an absolute error of 2.07. When we compared the absolute error of diversity estimation in different types of repertoires among T cell rich samples, all the methods offered more precise diversity estimation in monoclonal than in polyclonal samples. The average absolute error in T cell rich monoclonal samples was 0.54 while the average absolute error in T cell rich polyclonal repertoires was 4.05 (Fig.2d). Notably, MiXCR had the best performance in estimating diversity in monoclonal repertoires but the worst performance in polyclonal repertoires (Fig.2d). Among the T cell poor monoclonal samples, the average absolute error was 2.47 across the surveyed methods, which was larger than in T cell rich monoclonal samples (Fig.2d). We further investigated whether tissue types influence the estimates of diversity based on polyclonal samples, as we had polyclonal samples in all five tissue types. On average, MiXCR and ImReP had the smallest absolute errors in lymph nodes while TRUST4 has the smallest absolute error in kidney samples (Supplementary Fig.6a).

We also calculated clonality to account for the substantial differences in the number of clonotypes which the Shannon Diversity Index doesn’t account for directly. All these methods provided precise estimates of clonality for monoclonal repertoires and polyclonal repertoires of T cell rich tissues with an average absolute error of 0.0635 and 0.05598, respectively (Supplementary Fig.6b). Among the surveyed methods, MiXCR provided the most precise estimates for both clonality and diversity in T cell rich monoclonal repertoires. Even though these methods captured much fewer clonotypes compared to TCR-Seq results in T cell rich polyclonal repertoires, if the clonotype counts were taken into account by calculating clonality, these methods were able to offer comparable diversity estimates in T cell rich polyclonal repertoires as they were in T cell rich monoclonal repertoires.

### RNA-Seq based TCR profiling methods were able to successfully estimate relative frequencies of clonotypes in monoclonal repertoires

While differences in diversity and clonality estimates were observed, we further examined whether the surveyed methods were able to provide accurate estimates of the frequency of assembled clonotypes in different types of repertoires. The results showed that all three RNA-Seq based methods accurately estimated the relative frequencies in monoclonal repertoires (Fig.3a-f), especially in T cell rich tissues (MiXCR: r=0.9998, *p*<0.001; ImReP: r=0.9977, *p*<0.001; TRUST4: r=0.9985, *p*<0.001). The clonotype frequency between results from TCR-Seq and from RNA-Seq were positively correlated in T cell rich polyclonal repertoires (MiXCR: r=0.7715, *p*=3.1e-77; ImReP: r=0.6495, *p*=5.3e-240; TRUST4: r=0.6762, *p*=4.6e-281) (Fig.3g-i).

**Figure 3.**
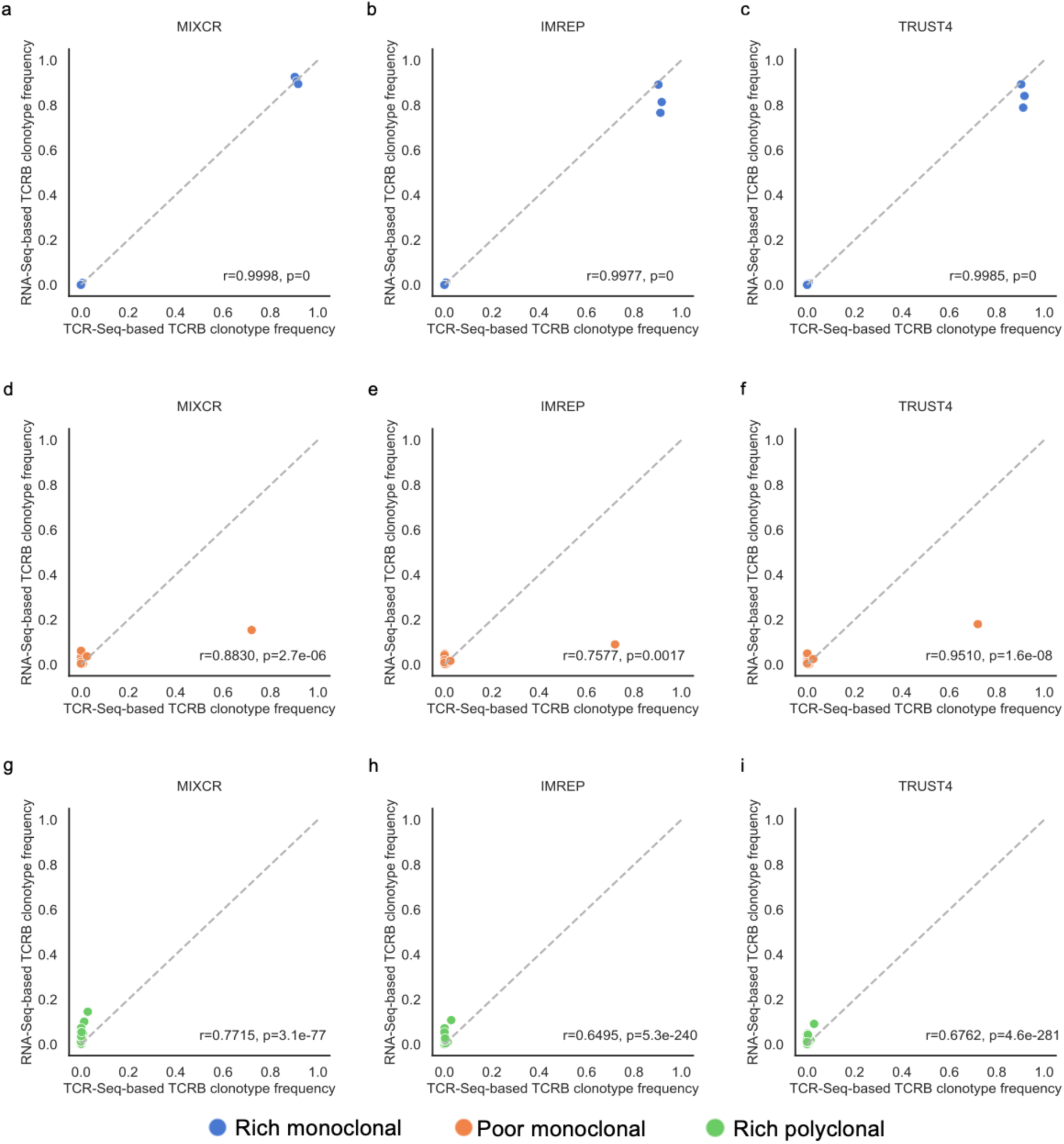
Correlation of TCRβ clonotype frequency based on the TCR-Seq data (x-axis) and the TCR derived reads from RNA-seq (y-axis). Only clonotypes assembled from RNA-Seq data are presented. Pearson correlation coefficients and the corresponding p-values are calculated and reported. a-c. The results of T cell rich monoclonal samples. d-f. The results of T cell poor monoclonal samples. g-i. The results of T cell rich polyclonal samples.

These findings suggested that all surveyed RNA-Seq-based methods were able to provide accurate estimations of relative frequencies of the detected clonotypes in monoclonal repertoires, while the positive correlations were weaker in polyclonal repertoires. MiXCR provided the best results in correlation of clonotypes in T cell rich polyclonal repertoires than those provided by ImReP and TRUST4.

### Fewer clonotypes were detected by RNA-Seq-based TCR profiling methods compared to TCR-Seq-based methods

Next, we compared the number of clonotypes detected by RNA-Seq-based methods compared to TCR-Seq-based methods. On average, TRUST4 captured the most clonotypes among three RNA-Seq-based methods with 4,451 in T cell rich monoclonal repertoire and 1,038 in T cell rich polyclonal repertoire, while MiXCR had the lowest with 1,723 in T cell rich monoclonal repertoire and 145 in T cell rich polyclonal repertoire (Supplementary Fig.7a). When we considered the absolute count of the TCR-Seq confirmed clonotypes, TRUST4 had the largest number of clonotypes confirmed by TCR-Seq, while MiXCR had the lowest (Supplementary Fig.7b). For example, on average, TRUST4 detected 538 TCR-Seq confirmed clonotypes and MiXCR only detected 317 in T cell rich monoclonal repertoires. These TCR-Seq confirmed clonotypes were highly overlapped across the survey methods in monoclonal repertoires (Supplementary Fig.7c-d). However, when we considered the fraction of the clonotypes that was confirmed by TCR-Seq in T cell rich tissues, MiXCR had the highest percentage of TCR-Seq confirmed clonotypes among all the TCR clonotypes that MiXCR detected, while ImRep and TRUST4 had similar performance (Supplementary Fig.7e). Combined with our findings above, these results suggested that ImRep and TRUST4 were able to detect more clonotypes including clonotypes with low frequencies than MiXCR. However, ImRep and TRUST4 had a higher portion of the detected clonotypes that were not confirmed by TCR-Seq than MiXCR in T cell rich tissues.

### The effect of the number of TCR derived reads on the performance of RNA-Seq-based TCR profiling methods

We examined how the number of receptor derived reads from RNA-Seq data influences the diversity estimates and capturing ability of the RNA-Seq-based methods. TCR derived reads were calculated by the total number of TCR reads matching the assembled clonotypes detected by RNA-Seq-based TCR profiling methods. To investigate the effect of TCR derived reads, we subsampled the RNA-Seq TCR derived reads from the original to 310, which 310 is the average TCR derived reads in T cell poor tissues, based on the original RNA-Seq assembled TCR clonotype frequencies from surveyed methods in three T cell rich monoclonal samples. We first measured the ability of RNA-Seq-based methods to estimate the diversity among these computationally modified samples by using the Shannon Diversity Index (SDI). The results indicated an increased number of TCR derived reads resulted in a more precise diversity estimate (Supplementary Fig.8a). With the increase of TCR derived reads, ImReP achieved the highest precision in estimating diversity. For example, ImReP had an absolute error of 0.26 in estimating SDI when the number of TCR derived reads was 28,000 (Supplementary Fig.8a).

We then investigated the portion of repertoire captured by these methods depending on the number of TCR derived reads presented in the sample. As the number of receptor derived reads decreases, the methods were more likely to miss the clonotypes with low abundance. For example, all TCRβ clonotypes with frequencies above 0.094% were captured across all three surveyed methods when the number of TCR derived read was 310 (Supplementary Fig.8b-d). While, without reducing TCR derived reads, all TCRβ clonotypes with frequencies greater than 0.036% were captured. The surveyed methods failed to detect low abundant clonotypes also contributed to the reduced accuracy in estimating diversity with the reduced number of TCR derived reads.

### The effect of read length on the performance of RNA-Seq-based TCR profiling methods

We investigated the effect of RNA-Seq read length on the ability of RNA-Seq-based methods to characterize the TCR repertoires by measuring the portion of repertoire that can be captured as well as diversity estimates. The length of RNA-Seq read was computationally reduced to 50bp and 75bp for PBMC samples.

RNA-Seq-based methods were able to capture the majority of the clonotypes in T cell rich monoclonal samples even with the reduced read length (Supplementary Fig.9a-c). These methods captured at least 91.8% of the clonotypes regardless of the read length that we examined in T cell rich monoclonal samples. All of the methods captured the entire repertoire with clonotype frequencies greater than 0.094% when the RNA-Seq samples had 50bp read length in T cell rich monoclonal samples, which this threshold was 0.036% with the original read length of 150bp. The portions of TCR clonotypes that can be captured were similar in T cell rich monoclonal repertoires among all read lengths, however, such portions dropped substantially with reduced read length in polyclonal samples (Supplementary Fig.9d-f). Only MiXCR captured all abundant clonotypes with reduced read lengths in T cell rich polyclonal samples.

Furthermore, when we examined the diversity estimate in the reduced read length samples, we noticed that the diversity estimates were only reliable in T cell rich monoclonal samples with reduced read length with an average of absolute error of 0.54 (the same value as the original read length) (Supplementary Fig.9g). In T cell rich polyclonal samples, the accuracy of estimating diversity was worse. The absolute error decreased with an increased RNA-Seq read length in ImReP and TRUST4, while the accuracy was similar between MiXCR results from 75bp and 150bp (Supplementary Fig.9h). RNA-Seq-based TCR profiling methods provided reliable results in T cell rich monoclonal samples across a variety of sequencing parameters.

### Performance of RNA-Seq methods in T cell poor tissues

In contrast to T cell rich monoclonal and polyclonal repertoires, all RNA-Seq based TCR profiling methods showed poor performance in T cell poor tissues. All the survey methods were not able to provide accurate diversity estimates in such tissues (MiXCR: r=0.2703, *p*=0.48; ImReP: r=0.3308, *p*=0.27; TRUST4: r=0.3073, *p*=0.31) (Supplementary Fig.10). When comparing the clonotype frequencies, the positive correlations for T cell poor polyclonal repertoires were weaker compared to other tissue and repertoire types (MiXCR: r=0.5165, *p*=3.2e-12, ImRep: r=0.2840, *p*=4.2e-07, TRUST4: r=0.6466, *p*=1.4e-61) (Supplementary Fig.11).

### Complement the TCRβ chain analysis with TCRα chain analysis

TCR-Seq requires separate library preparation and sequencing for α and β chains, while RNA-Seq can capture all four TCR chains (α, β, γ and δ chains) at the same time with single library preparation and sequencing. We compared the results for the TCRα chain with the TCRβ chain across different tissues and repertoire types. However, we have determined that delta and gamma chains did not have enough reads to be reliably profiled.

First, we compared the number of TCRα and TCRβ derived reads from RNA-Seq. The average number of TCR α chain reads derived from RNA-Seq were higher than the number of TCR β chain reads derived from RNA-Seq in T cell rich monoclonal repertoires, while more reads from β chains than α chains in T cell rich polyclonal repertoires (Supplementary Fig.12a-b). The number of TCRα and TCRβ derived reads were highly positively correlated in T cell rich tissues across all surveyed methods (MiXCR: r=0.9998, *p*=0.013; ImReP: r=0.9956, *p*=0.06; TRUST4: r=0.9855, *p*=0.11). The surveyed methods were still able to provide positive correlations in T cell poor polyclonal repertoires (MiXCR: r=0.9913, *p*=5e-11; ImReP: r=0.4072, *p*=0.17; TRUST4: r=0.9853, *p*=8.76e-10).

Next, we compared the number of TCRα and TCRβ clonotypes. Similar results were observed that more TCRα and TCRβ clonotypes were captured by RNA-Seq-based methods in monoclonal repertoires than polyclonal repertoires among T cell rich tissues (Supplementary Fig.12c-d). The number of clonotypes from α and β chains was comparably similar across repertoire types by RNA-Seq-based methods. All methods provided highly similar numbers of TCRα and TCRβ clonotypes in T cell rich tissues. The correlations were highly positive in T cell poor repertoires by MiXCR and TRUST4.

## Discussion

TCR-Seq is a powerful tool to profile TCR repertoires, gain novel insight into immunological phenomena, and serve as a biomarker for immunotherapy efficacy. However, such technology is cost-prohibitive in clinical cohorts, and is often unable to be routinely applied in such settings^19^. In contrast, RNA-seq is becoming a default technology to profile patient tissues in clinical pathology^20^. By utilizing an RNA-Seq approach to characterize the presence and relative frequency of TCR sequences within a given blood sample or T cell rich tissue sample, clinical samples can be more efficiently utilized without the need for additional dedicated sequencing experiments using TCR-Seq.

This study is the first one to benchmark the performance of RNA-Seq-based TCR profiling methods in both T cell rich and poor tissues across the largest number of clinical samples. While the previous study only included a certain type of TCR repertoire^16^, we expanded the examination to a broad range of cancer tissue types and repertoire types. We determined the scenarios under which RNA-Seq analysis using RNA-Seq-based TCR profiling methods can offer a comparable quality of characterizing immune repertoires. Given tissue with a sufficient degree of T cell level and with one or a few hyperexpanded clonotypes made up for the whole repertoires, and with adequate RNA sequencing depth and satisfactory quality, RNA-Seq-based TCR profiling methods are able to capture the majority of TCRβ clonotypes and effectively estimate the diversity of the repertoire. Despite the inability of RNA-Seq-based TCR profiling methods to capture the ultra-rare clonotypes supported by several TCR-Seq reads, these methods are generally able to capture the clonotypes with a frequency greater than 1.553% in T cell rich tissues. This suggests that RNA-Seq can potentially complement TCR-Seq technology in T cell rich tissues to successfully estimate the overall diversity of the sample and effectively detect clones with greater frequencies. The diversity estimated based on high-throughput measures usually underestimates true diversity^2,5^, but in cases where there is a major clonotype existing with high relative frequency than even capturing the small portion of the repertoire, one can estimate the diversity.

In more heterogeneous tissues which are more relatively T cell poor tissues, such as tumor biopsies, RNA-Seq-based TCR profiling methods are only able to capture the dominant clonotypes with the highest frequencies. This results in unreliable estimates of diversity and clonality in such tissues. These suggested that extra caution needs to be taken when utilizing RNA-Seq-based TCR profiling methods in T cell poor tissues, especially in polyclonal samples of the T cell poor tissues.

The ability to effectively characterize the TCR repertoire depends on the proportion of the total repertoire captured by a given technology. Even state-of-the-art high-throughput technologies fail to capture the entire diversity of the T cell repertoire; instead of capturing the most common clones as this is often sufficient to effectively characterize TCR diversity. Identification of dominant TCR clones within responding patient biopsies can be useful for the characterization of potential neoantigens being targeted by the TCRs. Capturing rare clones is extremely challenging due to the enormous diversity of the individual T cell repertoire and limited throughout of commercially available TCR-Seq protocols. As a result, rare clones can go unsampled and remain undetected in high-throughput measurements of the samples.

Diversity is measured based on the clonotypes that are present in the sequencing results and their relative frequencies. The surveyed methods are able to provide reliable diversity estimates in T cell rich monoclonal repertoires but not so in polyclonal repertoires. As expected, the number of clonotypes detected by RNA-Seq-based methods is always lower than those from TCR-Seq. Clonotypes with low frequencies are not captured by the RNA-Seq-based methods, this may have caused the error in estimating diversity. In addition to the undetected clonotypes with low abundance, the differences in clonotype frequency estimation as well as in the number of detected clonotypes could be the reasons that cause differences in the accuracy of diversity estimates between various tissue types and repertoire types.

There are a few limitations of our study. First, there is no standardized method to distinguish monoclonal and polyclonal TCR repertoires. We chose the Shannon Diversity Index of value two as the threshold which represents the distribution of clonotypes in the samples that we used in the study. Second, TCR-Seq protocols that were used were not consistent across different study cohorts. The lymph node and small intestine samples were sequenced by rapid amplification of 5’ complementary ends (5’RACE) approach, while all other samples were sequenced by immunoSEQ (Adaptive Biotechnology, Seattle, WA). The biases across multiplex PCR and 5’RACE are known^21^, however, the differences between immunoSEQ and other TCR-Seq profiling methods were unknown. Third, TCR-Seq computational methods used for processing were different across study cohorts as well. TCR-Seq raw data from lymph node and small intestine samples were processed through MiXCR while the others were processed via internal methods at Adaptive Biotechnology. Unfortunately, Adaptive Biotechnology does not share raw TCR-Seq data or bioinformatics methods, so we were not able to process all samples through the same bioinformatics software. The use of MiXCR for processing both TCR-Seq and RNA-Seq on lymph node and small intestine samples may lead to biases in the results. Last, TCR derived reads corresponded to T cell levels. However, RNA-Seq-based TCR profiling methods may also detect natural killer (NK) cells. Though NK cells were relatively rare compared to T cells, they could be added to the number of T cells that were deduced from the TCR reads.

While available approaches require separate assays for the detection of the α and β chains of the TCR, RNA-Seq-based TCR profiling methods are able to detect both α and β chains simultaneously from a single library preparation, reducing the overall cost of the assay. With the increasing emphasis on personalized medicine, there is an increasing demand for cost-effective and reproducible measures in order to comprehensively characterize cancer cells and profile immune cell populations. This technique can be employed in large-scale clinical cohorts. Further, detection of TCR sequences at the RNA level provides increased assurances that such sequences are transcriptionally active and more likely to be biologically relevant. It also provides opportunities to reuse the existing RNA-Seq data instead of initiating new TCR-Sequencing. Finally, RNA-Seq has already been used in clinical specimens provides an additional analysis tool for use on a variety of clinical samples for patients receiving immunotherapy for the treatment of cancer. Overall, RNA-Seq-based methods are appealing alternatives for profiling T cell receptor repertoires in T cell rich tissues and monoclonal samples when TCR-Seq is not yet available.

## Method

### Isolate genomic DNA and RNA

Genomic DNA and RNA from patient-derived transgenic NY-ESO-1 TCR PBMCs and normal control PBMCs^17^ were isolated with an AllPrep DNA/RNA isolation kit according to the manufacturer’s instructions (Qiagen). For patient melanoma biopsies treated with anti-PD-1 blockade^12^, genomic DNA was isolated from formalin-fixed, paraffin-embedded tumor biopsy shavings on an Anaprep 12 nucleic acid extraction platform (BioChain), while RNA was isolated from tumor samples which had been preserved in RNAlater (Qiagen) and stored at -80°C using an AllPrep RNA Isolation kit, as above. Melanin was then removed from visibly pigmented samples using a PCR Inhibitor Removal Kit (Zymo Research, Irvine, CA).

### Generate DNA-based TCR-Seq data

TCRβ alleles were sequenced at 100,000 reads by Adaptive Biotechnologies. Briefly, this process utilizes a synthetic immune repertoire, corresponding to every possible biological combination of Variable (V) and Joining (J) gene segments for each TCR locus, spiked into every sample at a known concentration. These inline controls enable rigorous quality assurance for every sample assayed and allow for correction of multiplex PCR amplification bias, providing an absolute quantitative measure of T cells containing the transgenic TCR relative to the other endogenous TCR clonotypes, with no difference in amplification efficiency^22^. Productive TCRβ sequences, i.e., those that could be translated into open reading frames and did not contain a stop codon, were reported.

### Analyze PBMC and melanoma biopsy TCR-Seq data

The assembled CDR3 sequences and corresponding VDJ recombinations were obtained from Adaptive Biotechnologies. We have only considered clones supported by at least two reads. Only full-length clonotypes (starting with C and ending F) were considered. Other clonotypes were filtered out.

### RNA-Seq of PBMC and melanoma biopsy samples

For both PBMC and melanoma biopsy samples, mRNA libraries were generated using the Kapa Stranded mRNA Kit (Roche) and were subjected to 150 bp paired-end sequencing (PBMCs) or 100 bp paired-end sequencing (melanoma biopsies) on a HiSeq 3000 platform.

### Download renal carcinoma clear cell samples

We used RNA-seq data from the TCGA that corresponds to three renal carcinoma clear cell samples sequenced by RNA-Seq and TCR-Seq. RNA-seq data are from Illumina HiSeq sequencing of 50-bp paired-end reads. We downloaded the mapped and unmapped reads in BAM format from the TCGA portal (TCGA-CZ-5463-01A, TCGA-CZ-5985-01A, TCGA-CZ-4862-01A). The corresponding TCR-Seq data was downloaded from immuneACCESS® portal (https://clients.adaptivebiotech.com/pub/Liu-2016-NatGenetics).

### Download ileocecal lymph node and small intestine samples

We downloaded the RNA-Seq data of one sample of melanoma specimens from the ileocecal lymph node (SPX6730, run: SRR5233639) and another sample of melanoma specimens from the small intestine (SPX8151, run:SRR5233637) from SRA (Accession: PRJNA371303). The corresponding TCR-Seq data was downloaded from https://figshare.com/articles/dataset/Antigen_receptor_repertoires_profiling_from_RNA-Seq/4620739?file=9787642.

### Run RNA-Seq-based repertoire profiling tools

RNA-Seq-based repertoire profiling tools were run using the directions provided with each of the respective tools. Wrappers were then prepared in order to run each of the respective tools as well as create standardized log files. When running the tools, we have chosen the most appropriate parameters. For MiXCR, we used the single analyze shotgun command. MiXCR accepts the raw fastq files as input for generating a report of the clonotypes present within the sample. Starting with the RNA sequence data, we mapped the reads onto the human chromosome to generate the bam file. Together with the bam file and the indexed bam file generated from samtools, we ran ImReP or TRUST4 to extract TCR clonotypes and corresponding supporting reads. The computational pipeline to compare RNA-Seq-based repertoire profiling methods with the gold standard is open source, free to use under the MIT license, and available at https://github.com/Mangul-Lab-USC/TCR-Seq_benchmarking_publication.

### Estimate TCR diversity and relative frequency

To estimate the diversity within each sample we only consider clonotypes that are supported by at least 2 reads. The relative frequency of each clonotype is then calculated as the number of supporting reads divided by the total number of reads supporting all clonotypes that occur at least twice.

Shannon Diversity Index:

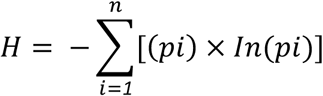

Clonality:

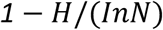

*pi* = frequency of clonotype i

*n* = number of unique clonotype in the sample

### Computational modify the properties of RNA-Seq samples

To reduce the length of RNA-Seq reads, we split each original sequence (150bp) from the raw fastq files into 2 and 3 segments and got new 75bp and 50bp sequences. The resulted sequences were written into new fastq files. We also computationally reduced the number of TCR derived reads in order to generate in silico samples based on the T cell rich monoclonal samples. The clonotypes that can be detected by the RNA-Seq based methods were randomly selected, in proportional to the original clonotype frequency in the samples. The total numbers of TCR derived reads from each tool were reduced to around 310, 1300, 2600, 13500, 28000.

## Data Availability

RNA-Seq data for five PBMC samples are available in Sequence Read Archive (SRA) under BioProject ID: PRJNA812076.

## Code availability

The code that was used to produce the figures and analysis performed in this paper are available at https://github.com/Mangul-Lab-USC/TCR-Seq_benchmarking_publication.

## Data availability

SRA data was downloaded via the SRA archive (https://www.ncbi.nlm.nih.gov/sra). RNA-Seq data for five PBMC samples are available in Sequence Read Archive (SRA) under BioProject ID: PRJNA812076. All processed and summarized data required to produce the figures and analysis performed in this paper are available at https://github.com/Mangul-Lab-USC/TCR-Seq_benchmarking_publication, including the data used to produce all figures.

## Acknowledgments

KP and SM are supported by the National Science Foundation grants 2041984 and 2135954. TSN is supported by the NIH grant K08 CA241088 and the Hyundai Hope on Wheels Hope Scholar Award. The authors acknowledge the Center for Advanced Research Computing (CARC) at the University of Southern California for providing computing resources that have contributed to the research results reported within this publication. URL: https://carc.usc.edu.

## Supplementary Tables

**Supplementary Table 1.** Overview of the samples in the gold standard dataset. The samples are sorted by the sample name (indicated in the column “Sample name”). We documented the tissue of which the sample was taken (indicated in the column “Tissue”), the tissue type based on the T cell levels (indicated in the column “Tissue type”), the repertoire type based on the Shannon Diversity Index (indicated in the column “Repertoire type”), the number of TCR-Seq reads from TCR-Seq data (indicated in the column “Number of TCR-Seq reads”), the number of RNA-Seq reads from RNA-Seq data (indicated in the column “Number of RNA-Seq reads”), the number of derived reads from RNA-Seq data after processing through RNA-Seq-based TCR profiling methods (indicated in the columns “Number of derived reads from RNA-Seq (MiXCR)”, Number of derived reads from RNA-Seq (ImReP), and Number of derived reads from RNA-Seq (TRUST4), respectively).

| No. | Sample name | Tissue | Tissue type (T cell rich or poor) | Repertoire type | Number of TCR-Seq reads | Number of RNA-Seq reads | Number of derived reads from RNA-Seq (MiXCR) | Number of derived reads from RNA-Seq (ImReP) | Number of derived reads from RNA-Seq (TRUST4) |
| --- | --- | --- | --- | --- | --- | --- | --- | --- | --- |
| 1 | sample01 | PBMC | T cell rich | monoclonal | 90,577 | 104,984,482 | 132,472 | 219,185 | 264,884 |
| 2 | sample02 | PBMC | T cell rich | monoclonal | 87,762 | 73,892,845 | 41,370 | 57,700 | 89,947 |
| 3 | sample03 | PBMC | T cell rich | monoclonal | 305,953 | 71,654,976 | 52,066 | 64,914 | 99,723 |
| 4 | sample04 | PBMC | T cell rich | polyclonal | 104,779 | 43,179,989 | 158 | 1354 | 2006 |
| 5 | sample05 | PBMC | T cell rich | polyclonal | 18,617 | 40,524,817 | 55 | 111 | 186 |
| 6 | sample06 | melanoma | T cell poor | polyclonal | 906,121 | 82,476,159 | 91 | 231 | 689 |
| 7 | sample07 | melanoma | T cell poor | polyclonal | 1,257,571 | 72,397,468 | 0 | 2 | 19 |
| 8 | sample08 | melanoma | T cell poor | polyclonal | 1,408,590 | 85,492,431 | 150 | 259 | 657 |
| 9 | sample09 | melanoma | T cell poor | polyclonal | 1,157,845 | 68,584,414 | 310 | 334 | 1009 |
| 10 | sample10 | melanoma | T cell poor | polyclonal | 1,769,522 | 63,320,771 | 59 | 71 | 306 |
| 11 | sample11 | melanoma | T cell poor | polyclonal | 1,006,220 | 55,727,841 | 12 | 31 | 63 |
| 12 | sample12 | melanoma | T cell poor | polyclonal | 292,828 | 107,919,183 | 8 | 10 | 39 |
| 13 | sample13 | melanoma | T cell poor | monoclonal | 12,954 | 80,622,502 | 1071 | 844 | 2855 |
| 14 | sample14 | melanoma | T cell poor | polyclonal | 749,686 | 55,931,661 | 113 | 152 | 351 |
| 15 | SRR5233637 | small intestine | T cell poor | polyclonal | 3,047,629 | 72,720,367 | 84 | 315 | 544 |
| 16 | SRR5233639 | lymph node | T cell rich | polyclonal | 3,256,697 | 65,998,918 | 1346 | 6143 | 9054 |
| 17 | TCGA-CZ-4862 | renal cell | T cell poor | polyclonal | 16,784 | 58,529,923 | 0 | 50 | 710 |
| 18 | TCGA-CZ-5463 | renal cell | T cell poor | polyclonal | 806 | 122,632,451 | 0 | 16 | 366 |
| 19 | TCGA-CZ-5985 | renal cell | T cell poor | polyclonal | 12,998 | 83,862,519 | 0 | 23 | 282 |

**Supplementary Table 2.** Execution of RNA-Seq-based TCR profiling methods. We documented the required modules for running the methods (indicated in the column “Load module”), the command used to process the RNA-Seq data (indicated in the column “Execute”), the link to the manual for methods (indicated in the column “Manual”).

| RNA-Seq-based methods | Load module | Execute | Manual |
| --- | --- | --- | --- |
| MiXCR | module load gcc/8.3.0<br>module load<br>openjdk/11.0.2 | /your/path/mixcr-3.0.13/mixcr analyze shotgun --species hs --starting-material rna --only-productive sample_1.fastq sample_2.fastq sample | https://github.com/mandricigor/ImReP/wiki |
| ImReP | module load gcc/4.9.4<br>module load<br>python/2.7.16 | python2<br>/your/path/ImReP/ImReP.py --extendedOutput --hg38 --bam --noOverlapStep --noCast sample.bam sample.cdr3 | https://mixcr.readthedocs.io/en/master/quickstart.html#id3 |
| TRUST4 | module load gcc/9.2.0<br>export<br>LC_CTYPE=en_US.UTF-8<br>export<br>LC_ALL=en_US.UTF-8 | /your/path/TRUST4/run-trust4 -b sample.bam -f hg38_bcrtcr.fa --ref human_IMGT+C.fa | https://github.com/liulab-dfci/TRUST4 |

**Supplementary Table 3.** Average number of TCRβ derived reads by RNA-Seq-based methods in different tissue types.

| Tissue | Average TCRβ derived reads (mean±SD) | | | |
| --- | --- | --- | --- | --- |
|  | MiXCR | ImReP | TRUST4 | Overall |
| PBMC | 45,224±54187 | 68,653±89,472 | 91,349±107,795 | 68,409±82,620 |
| Melanoma biopsies | 202±340 | 215±264 | 665±888 | 361±590 |
| Renal cell | 0 | 30±18 | 453±227 | 161±247 |
| Lymph node | 1346 | 6143 | 9054 | 5514±3892 |
| Small intestine | 84 | 315 | 544 | 314±230 |

**Supplementary Table 4.**
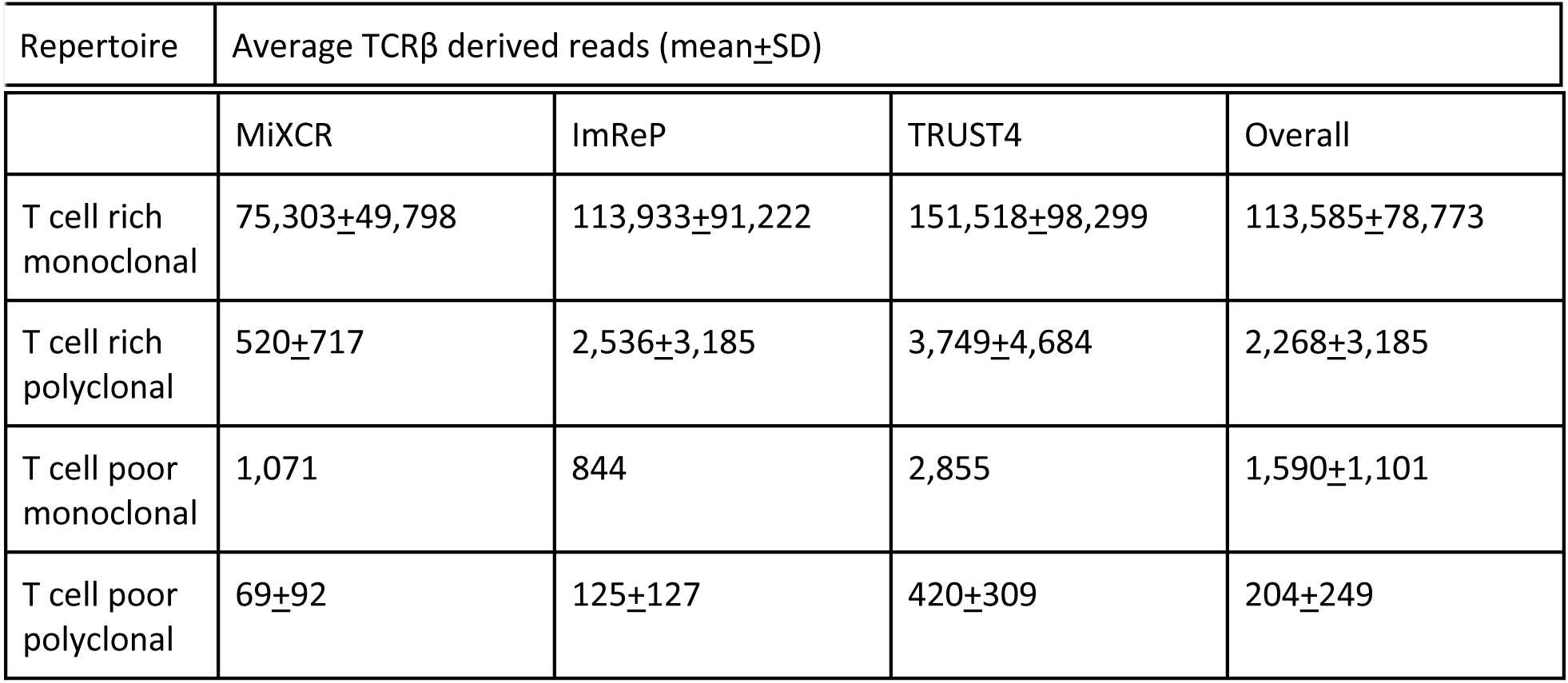
The average number of TCRβ derived reads by RNA-Seq-based methods in different repertoire types.

## Supplementary figures

**Supplementary Fig.1.**
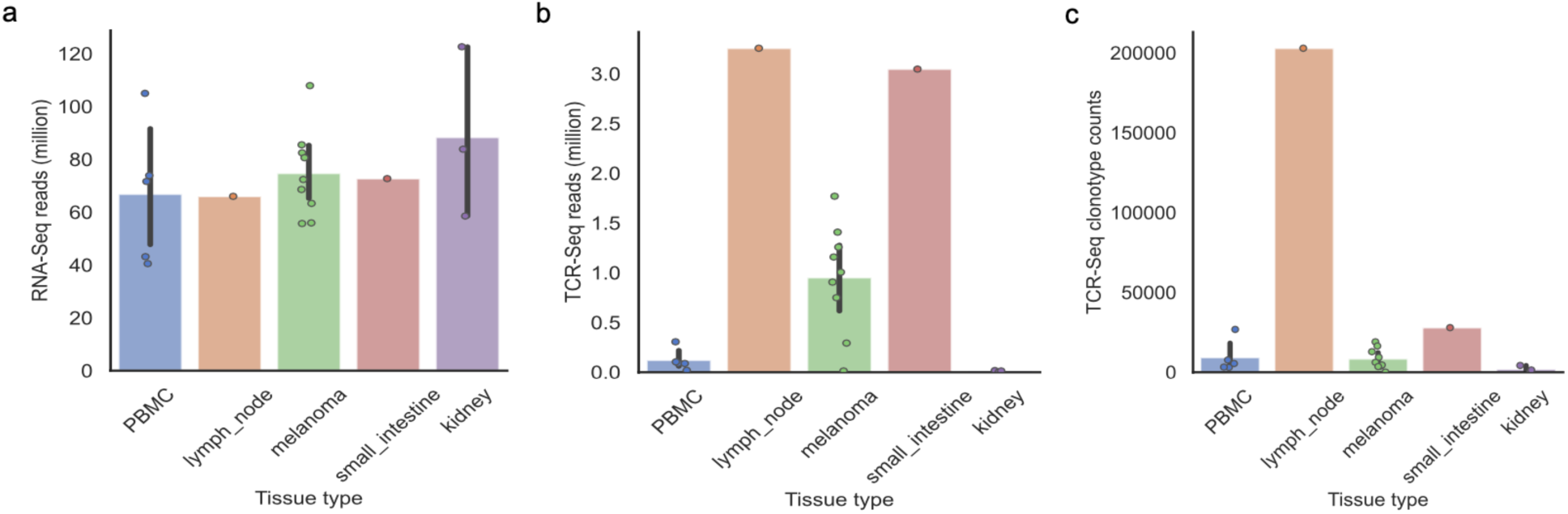
Basic metrics of the dataset. a. Bar plot and scatter plot of the number of RNA-Seq reads in million across different tissue types. b. Bar plot and scatter plot of the number of TCR-Seq reads in million across different tissue types. c. Bar plot and scatter plot of the number of TCRβ clonotype counts from TCR-Seq across different tissue types. PBMC samples are shown in blue, lymph node sample is shown in orange, melanoma samples are shown in green, small intestine sample is shown in red, kidney sample are shown in purple.

**Supplementary Fig.2.**
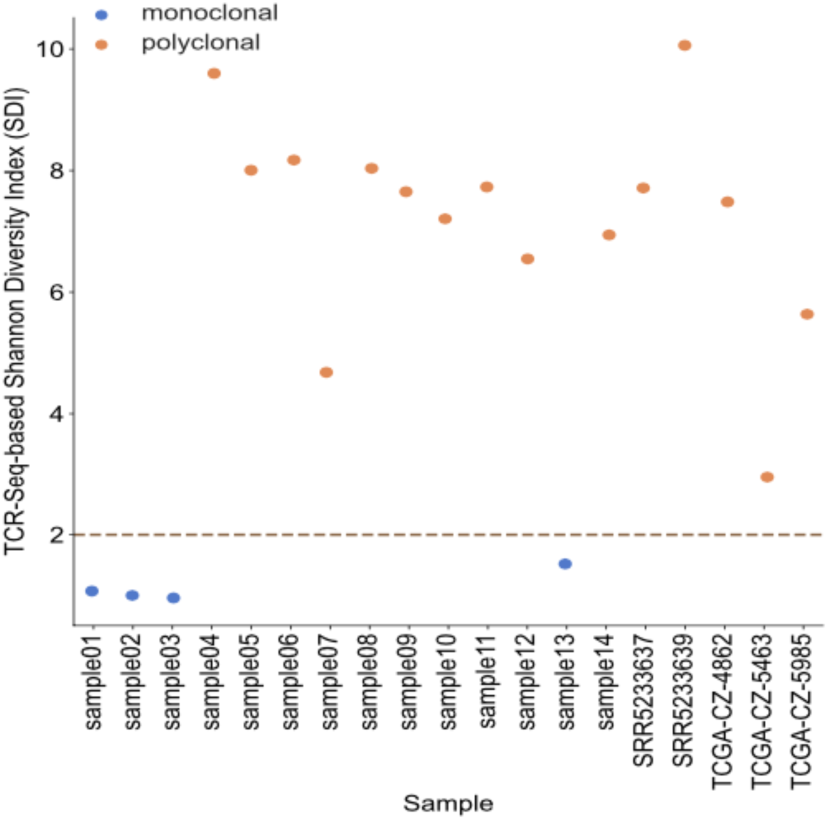
The Shannon Diversity Index (SDI) of each sample. Samples with SDI less than 2 are considered as monoclonal samples, otherwise polyclonal samples. The monoclonal samples are shown in blue and polyclonal samples are shown in orange.

**Supplementary Fig.3.**
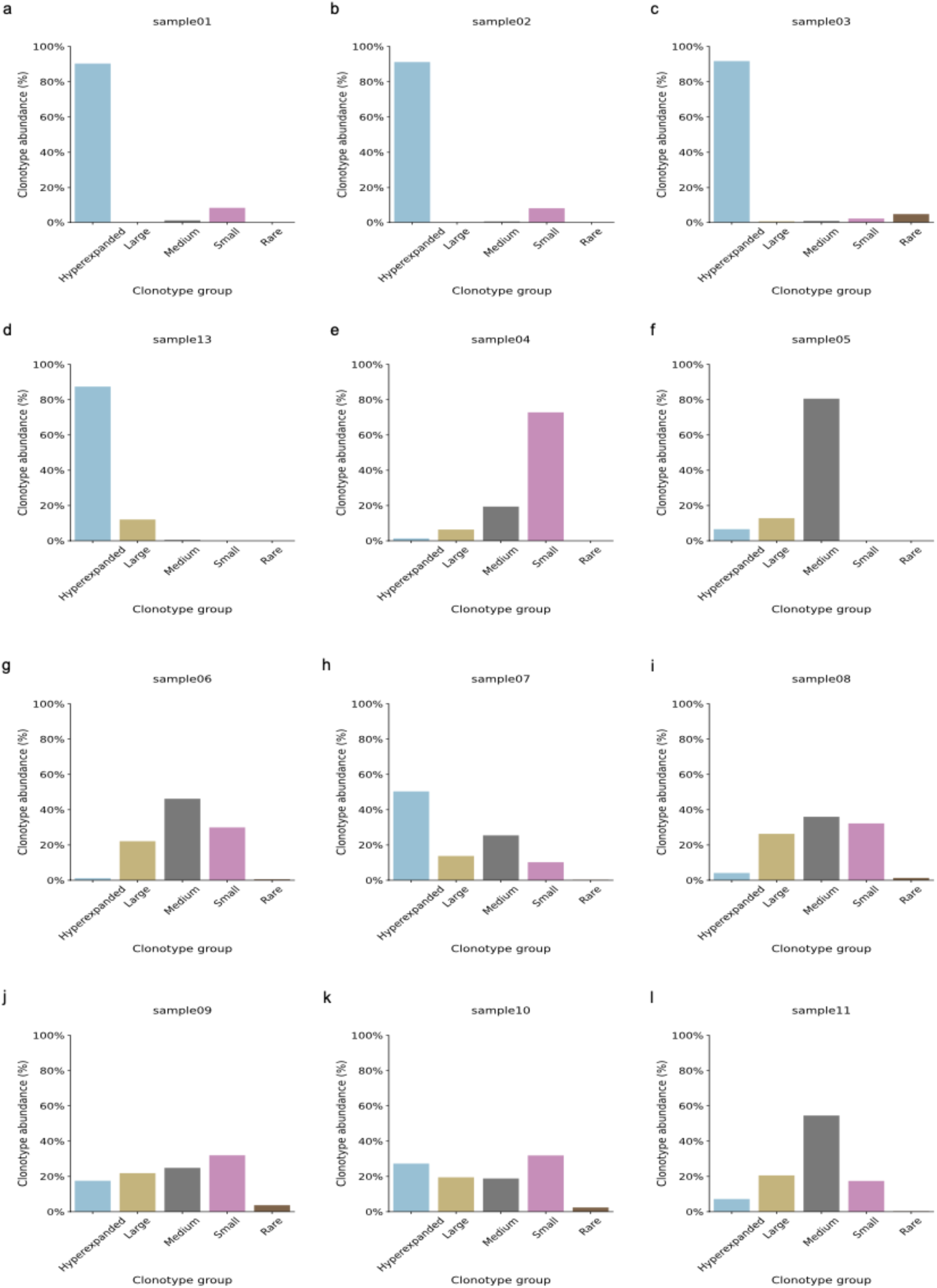
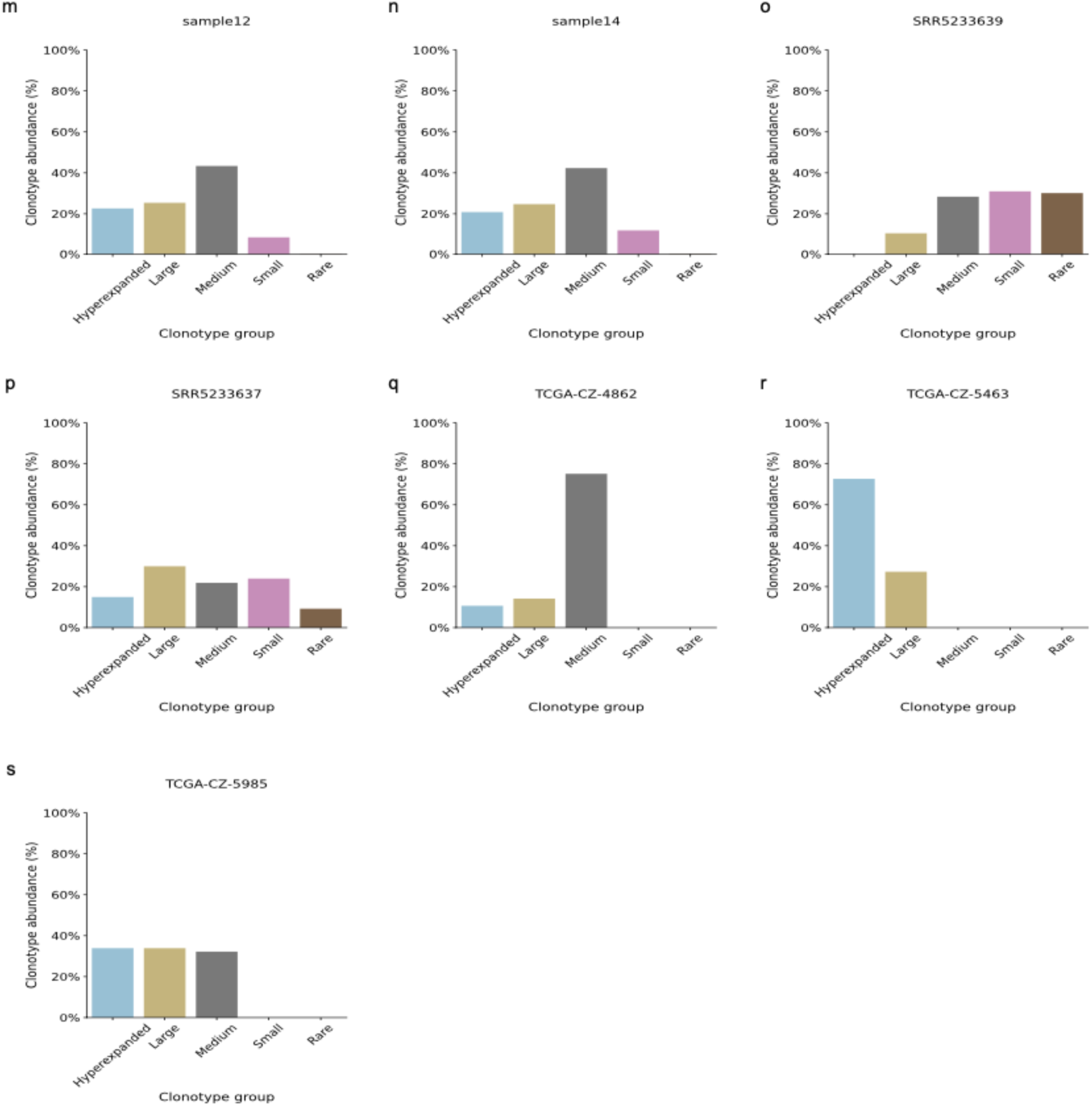
The distribution of clonotype groups in each sample. The clonotypes are categorized into five groups based on the clonotype frequencies. Hyperexpanded clonotypes (cyan) are the clones with frequencies between 0.01 to 1, large clonotypes (yellow) are the clones with frequencies between 0.001 to 0.01, medium clonotypes (gray) are the clones with frequencies between 0.0001 to 0.001, small clonotypes (magenta) are the clones with frequencies between 0.00001 to 0.0001, rare clonotypes (cambridge leather) are the clones with frequencies between 0 to 0.00001. a-d. The results of monoclonal samples. e-s. The results of polyclonal samples.

**Supplementary Fig.4.**
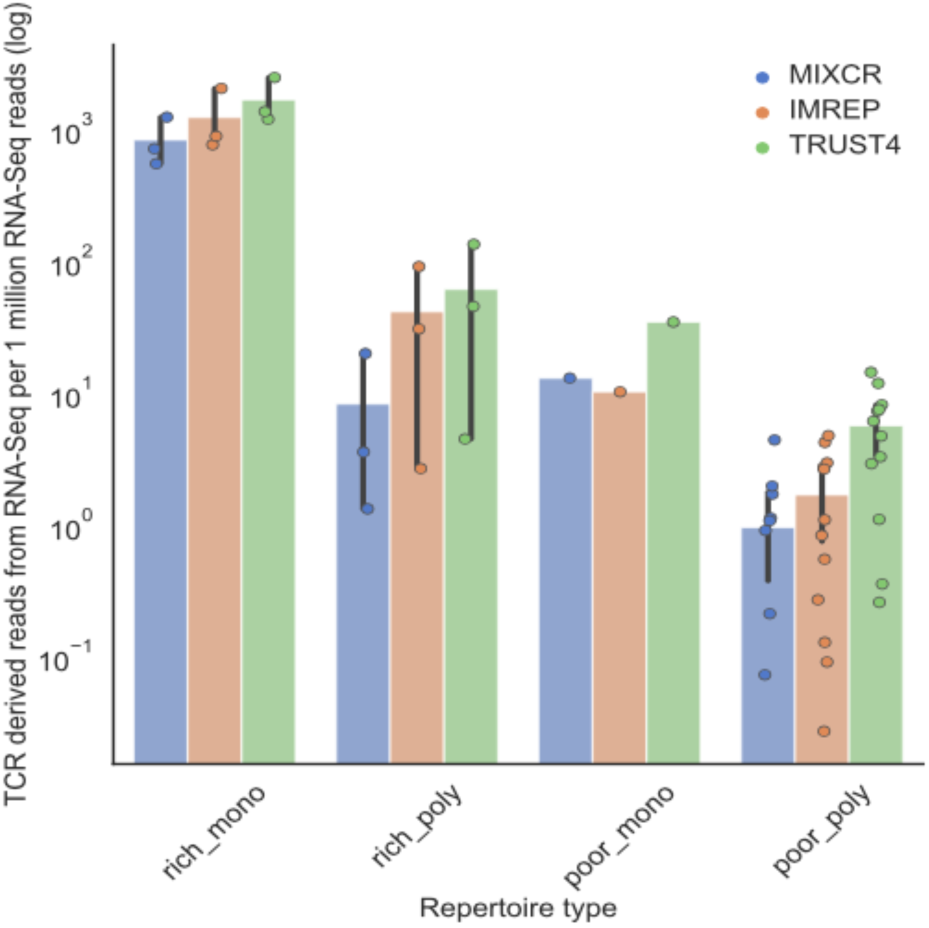
The number of TCR derived reads by RNA-Seq-based TCR profiling methods per 1 million RNA-Seq reads in T cell rich monoclonal samples (rich_mono), T cell rich polyclonal samples (rich_poly), T cell poor monoclonal samples (poor_mono), and T cell poor polyclonal samples (poor_poly). The y-axis corresponds to number of TCR derived reads on a log scale. The results from MiXCR are represented in blue, the results from ImReP are represented in orange, the results from TRUST4 are represented in green.

**Supplementary Fig.5.**
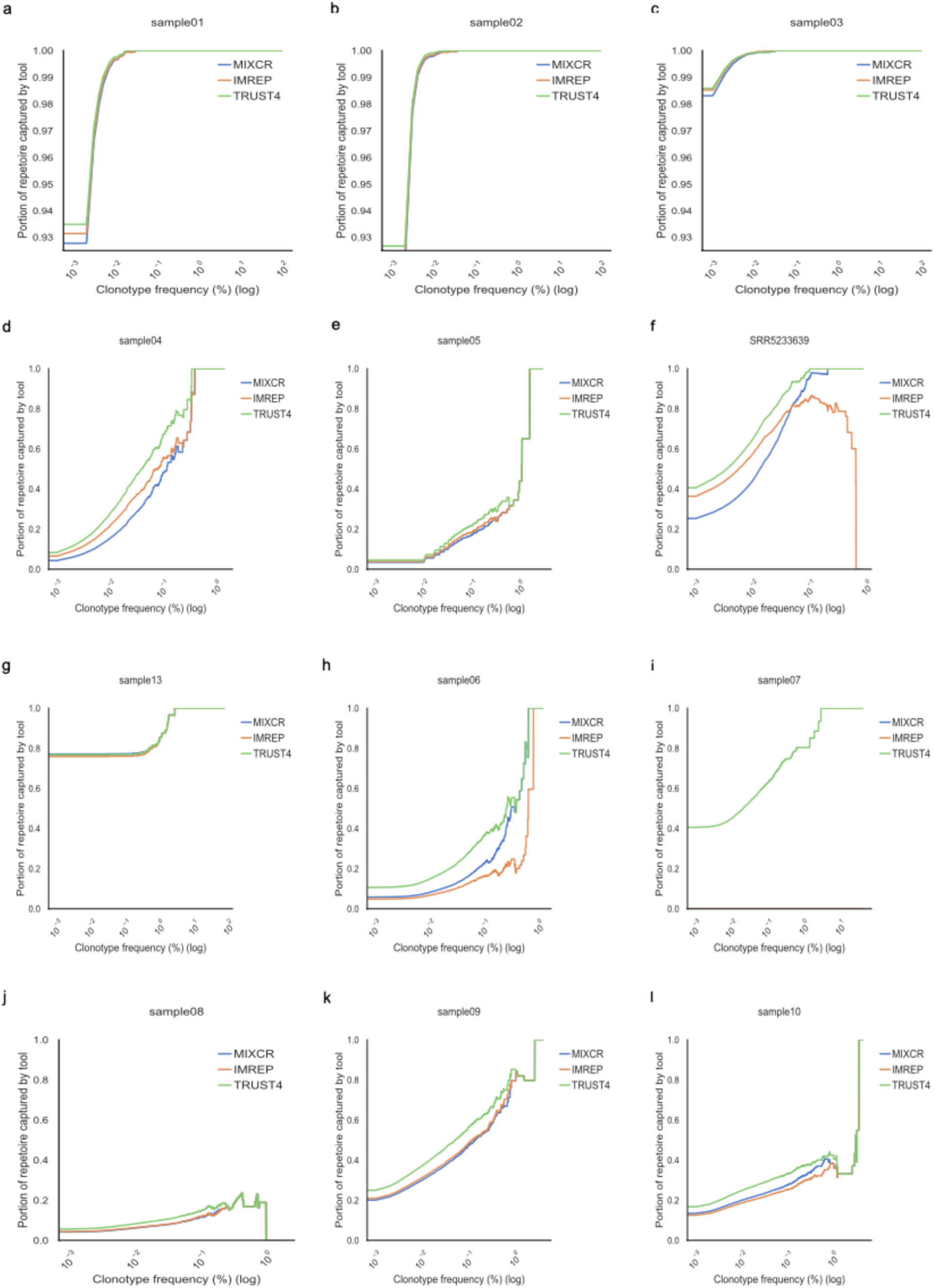
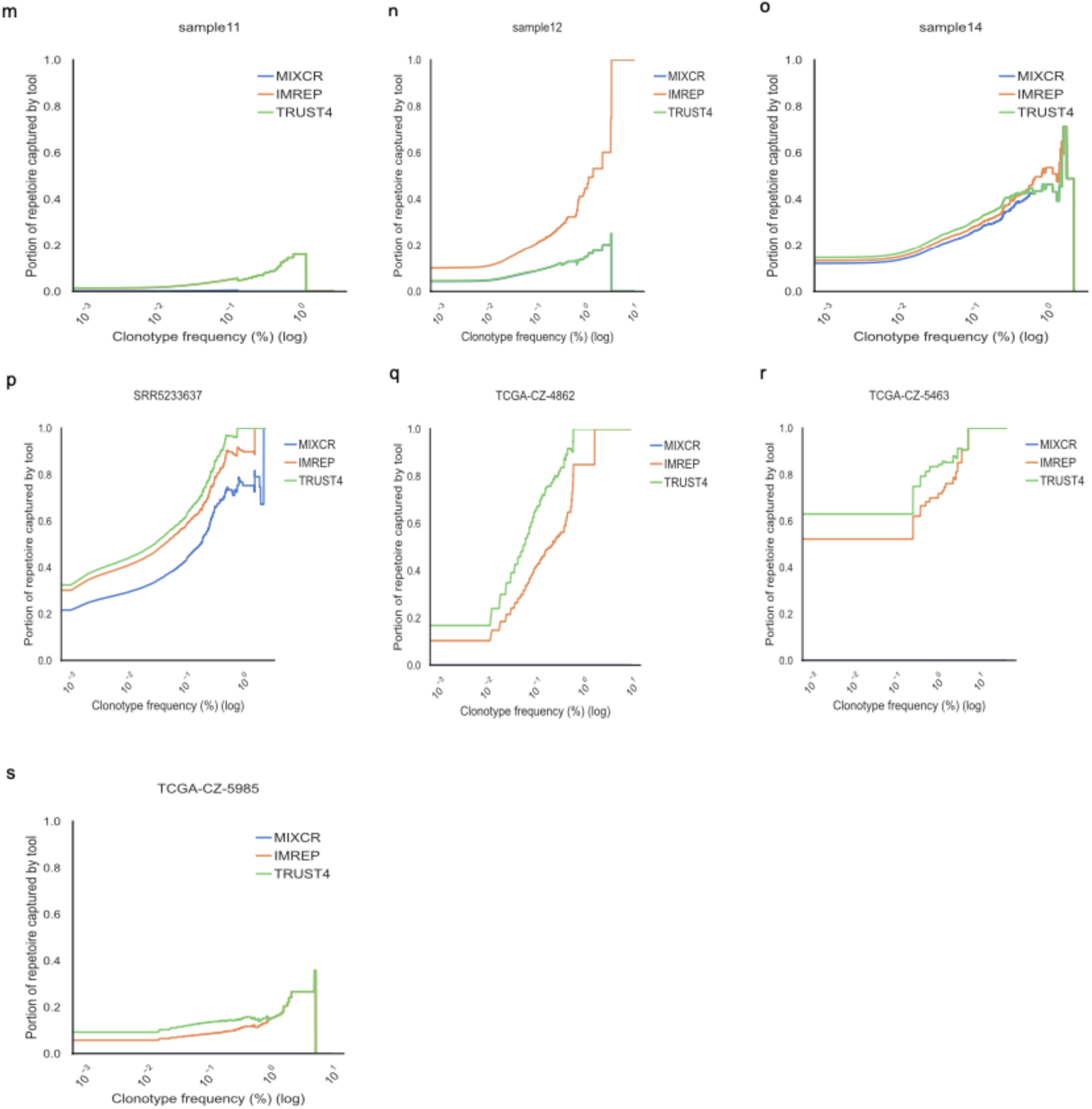
The capability of capturing TCR-Seq confirmed clonotypes by RNA-Seq-based methods in each sample. Line plots with x-axis corresponding to TCR-seq confirmed clonotypes with a frequency of Z, and y-axis corresponding to the fraction of assembled TCR repertoire by RNA-Seq-based methods with clonotype frequency greater than Z in (a-c) T cell rich monoclonal samples, (d-f) T cell rich polyclonal samples, (g) T cell poor monoclonal sample, and (h-s) T cell poor polyclonal samples. The capability of capturing by MiXCR, ImReP, and TRUST4 are represented in blue, orange, green, respectively.

**Supplementary Fig.6.**
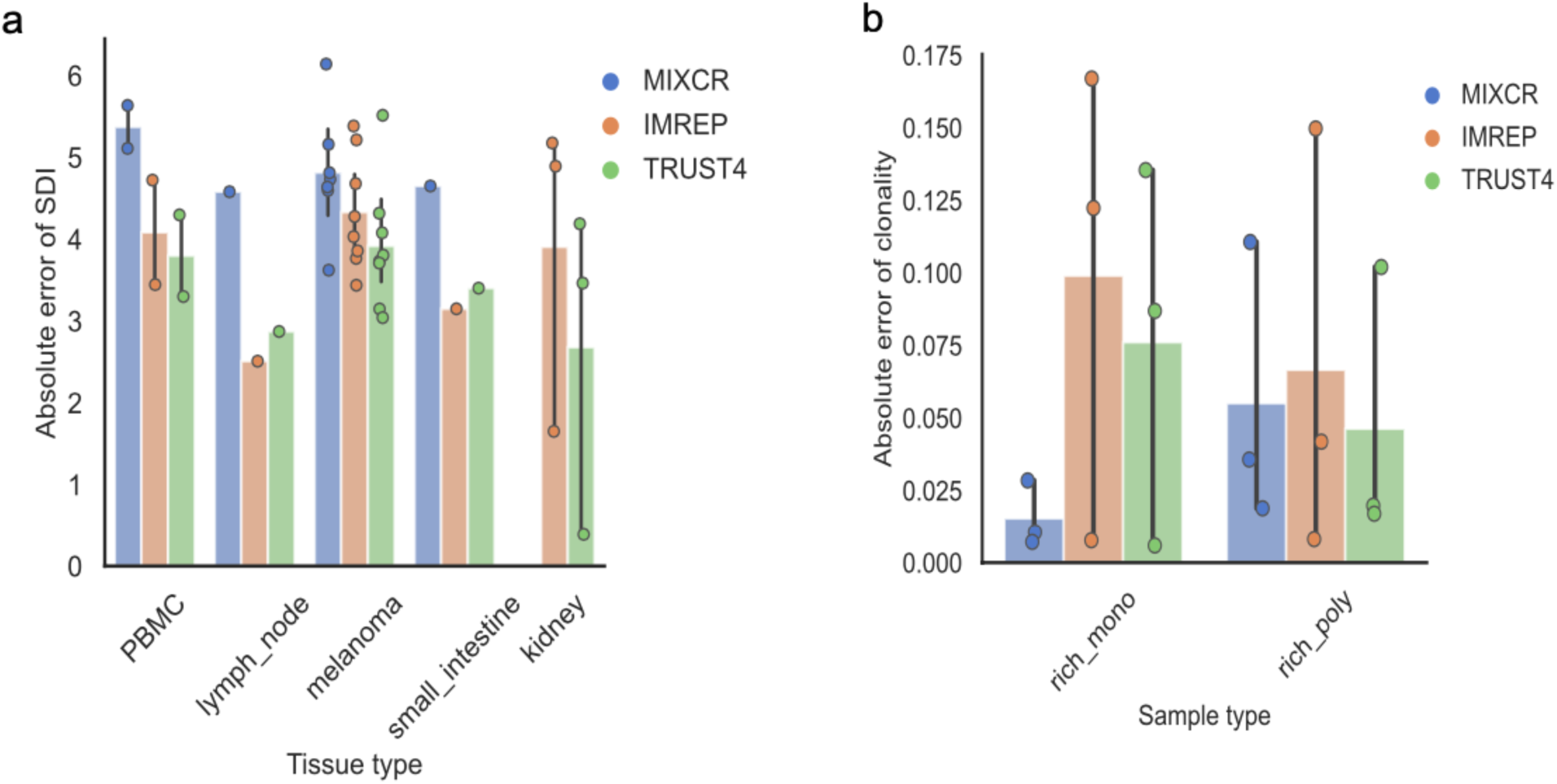
Diversity and clonality comparison between TCR-Seq-based results and RNA-Seq-based results. a. Absolute error between diversity estimated based on RNA-Seq data and TCR-Seq data by Shannon Diversity Index (SDI) among polyclonal samples in different types of tissues. b. Bar plot and scatter plot of absolute error between clonality estimated based on RNA-Seq data and TCR-Seq data in T cell rich monoclonal samples (rich_mono), and T cell rich polyclonal samples (rich_poly). The results from MiXCR are represented in blue, the results from ImReP are represented in orange, the results from TRUST4 are represented in green.

**Supplementary Fig.7.**
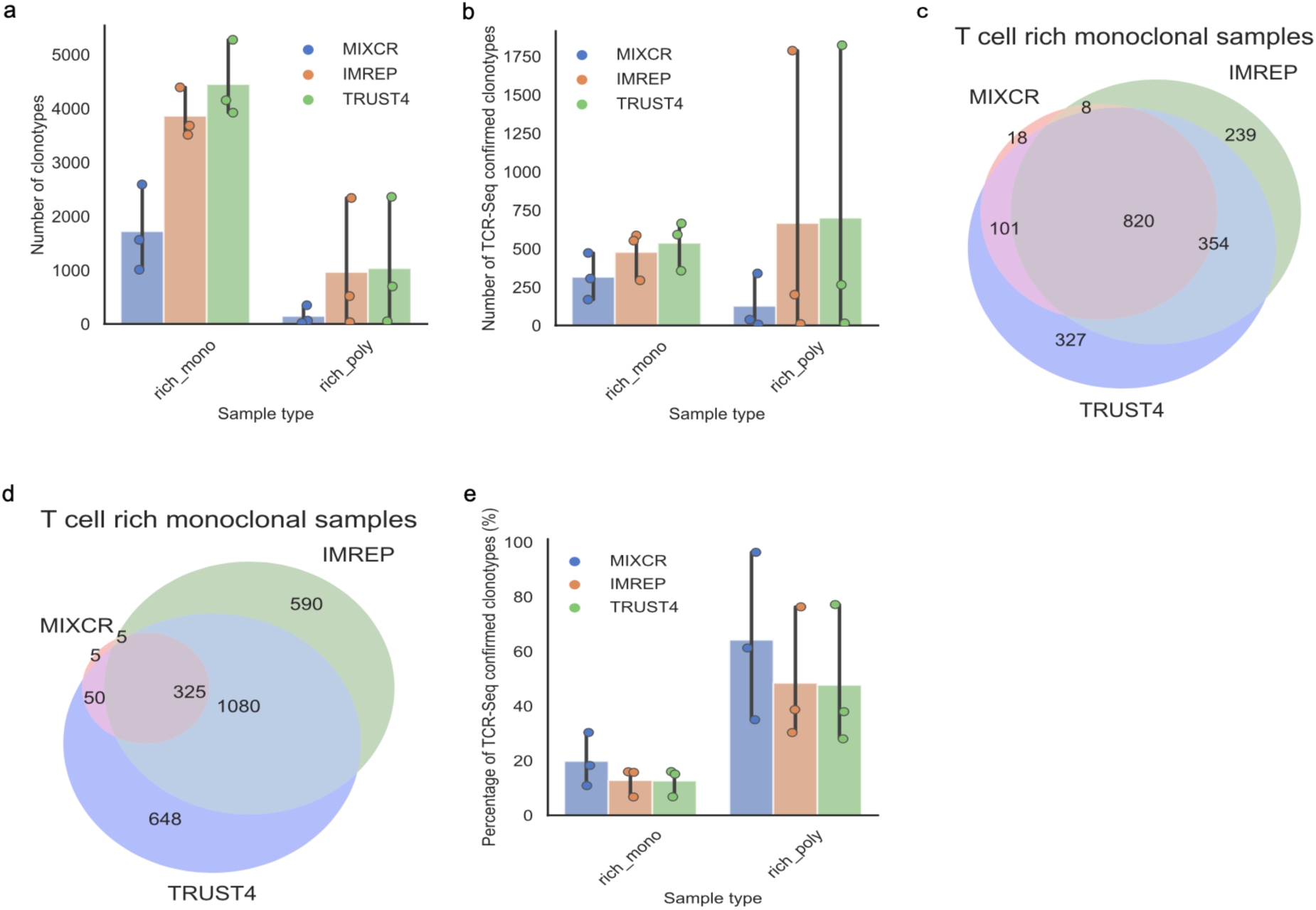
TCRβ derived clonotype counts from RNA-Seq data. a. Bar plot and scatter plot of the number of TCRβ derived clonotype counts in T cell rich monoclonal samples (rich_mono) and T cell rich polyclonal samples (rich_poly). b. Bar plot and scatter plot of the number of TCRβ derived clonotypes that are confirmed by TCR-Seq in T cell rich monoclonal samples (rich_mono) and T cell rich polyclonal samples (rich_poly). c-d. Venn diagrams of TCR-Seq confirmed clonotypes across MiXCR, ImReP, and TRUSTR4 in T cell rich tissues. The numbers indicate the sum of clonotype counts across three monoclonal samples (c) and four polyclonal samples (d). The overlapped numbers indicate the number of TCR-Seq confirmed clonotypes that are presented in results from two or three methods. e. Bar plot and scatter plot of the percentage of TCRβ derived clonotypes that are confirmed by TCR-Seq in T cell rich monoclonal samples (rich_mono) and T cell rich polyclonal samples (rich_poly). The results from MiXCR are represented in blue, the results from ImReP are represented in orange, and the results from TRUST4 are represented in green.

**Supplementary Fig.8.**
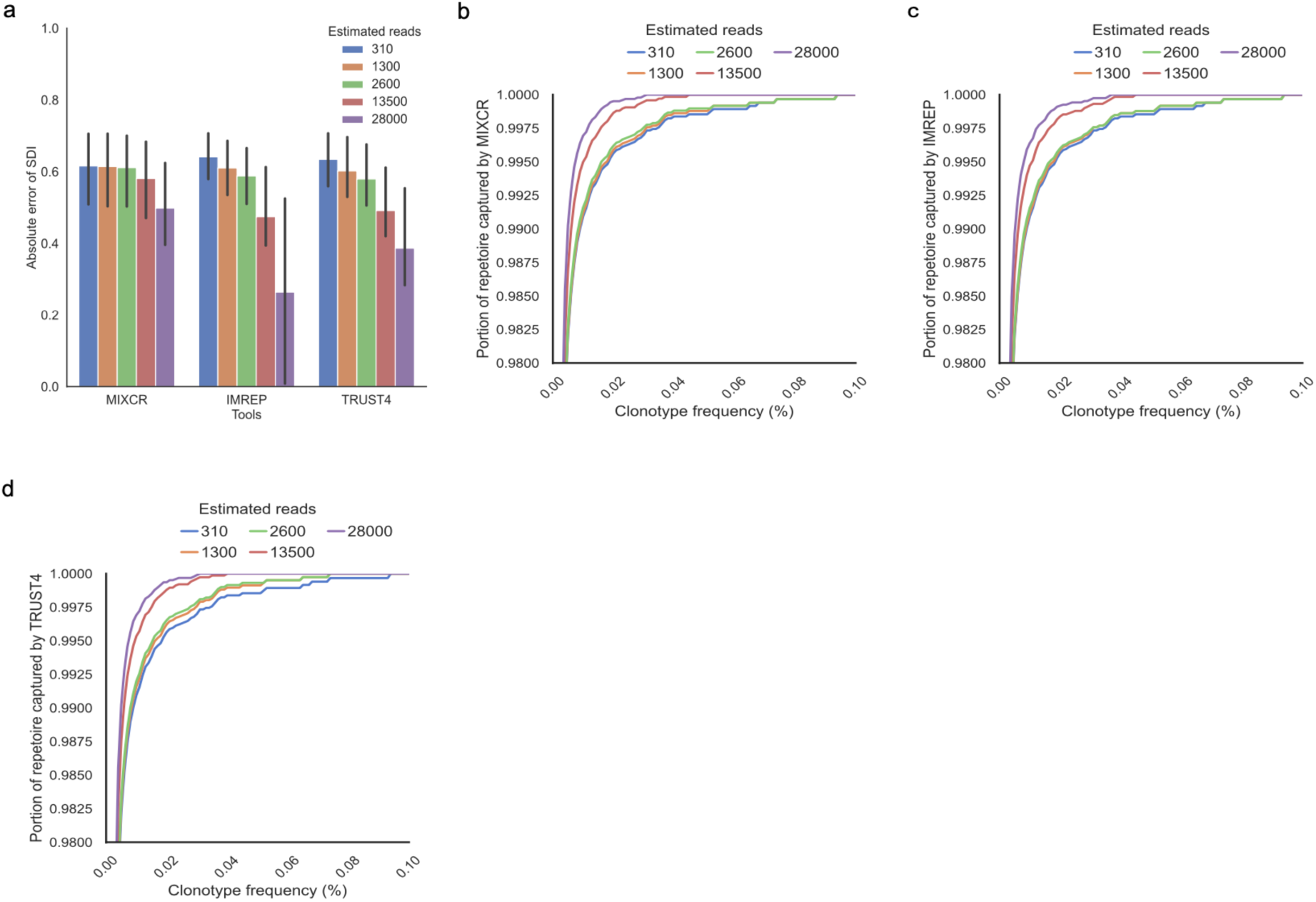
The absolute error of RNA-Seq based TCRβ diversity and TCR-Seq based TCRβ diversity and capability of capturing TCR-Seq confirmed clonotypes by RNA-Seq-based methods in computationally modified monoclonal repertoires with reduced reads. a. Bar plots that present the absolute error of SDI with respect to the total number of TCR derived reads across the RNA-Seq-based methods, with error bars with 95% confidence interval. b-d. Line plots with x-axis corresponding to TCR-seq confirmed clonotypes with a frequency of Z, and y-axis corresponding to the fraction of assembled TCR repertoire by RNA-Seq-based methods with clonotype frequency greater than Z. Results from estimated TCR derived reads of 310, 1300, 2600, 13500, and 28000 are presented in blue, orange, green, red, and purple, respectively.

**Supplementary Fig.9.**
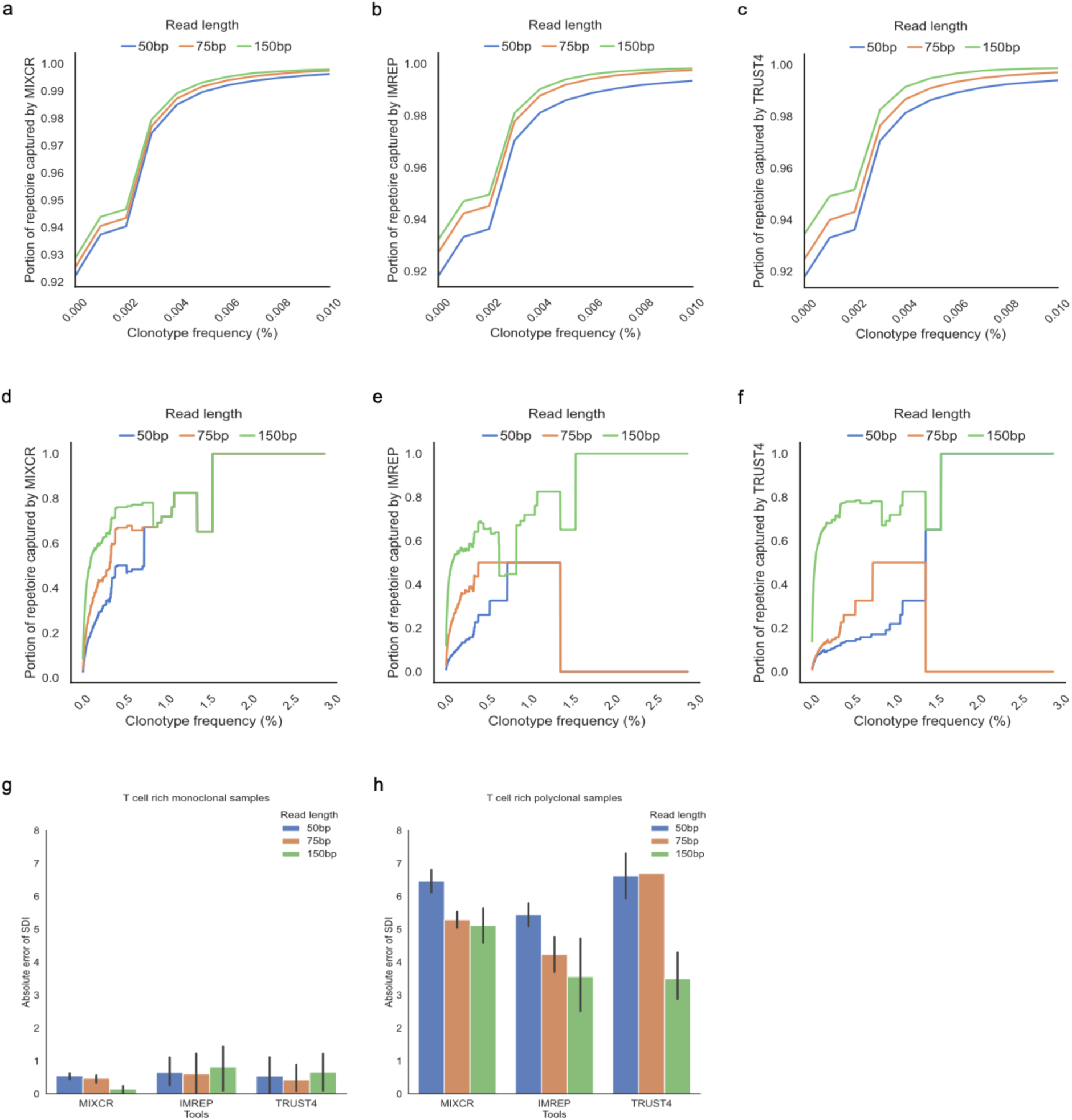
The capability of capturing TCR-Seq confirmed clonotypes by RNA-Seq-based methods and absolute error of RNA-Seq based TCRβ diversity and TCR-Seq based TCRβ diversity from reduced RNA-Seq read length results. a-f. Line plots with x-axis corresponding to TCR-seq confirmed clonotypes with a frequency of Z, and y-axis corresponding to the fraction of assembled TCR repertoire by RNA-Seq-based methods with clonotype frequency greater than Z in T cell rich monoclonal samples (a-c) and T cell rich polyclonal samples (d-f). g-h. Bar plots that present the absolute error of SDI with respect to RNA-Seq read length in T cell rich monoclonal samples and T cell rich polyclonal samples across the RNA-Seq-based methods, with error bars with 95% confidence interval. Results from RNA-Seq read length 50bp, 75bp, and 150bp are presented in blue, orange, and green, respectively.

**Supplementary Fig.10.**
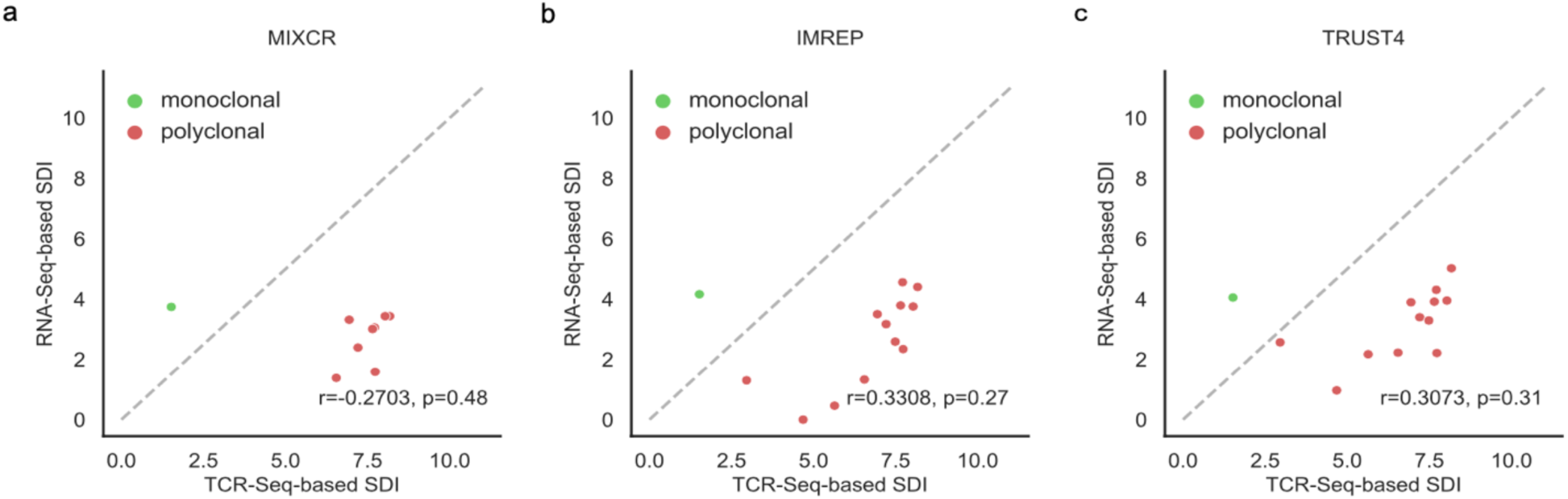
Correlation of diversity estimated based on RNA-Seq data (y-axis) and TCR-Seq data (x-axis) by Shannon Diversity Index (SDI) in T cell poor tissues. Green represents monoclonal samples, red represents polyclonal samples. Pearson correlation coefficients and the corresponding p-values are calculated and reported.

**Supplementary Fig.11.**
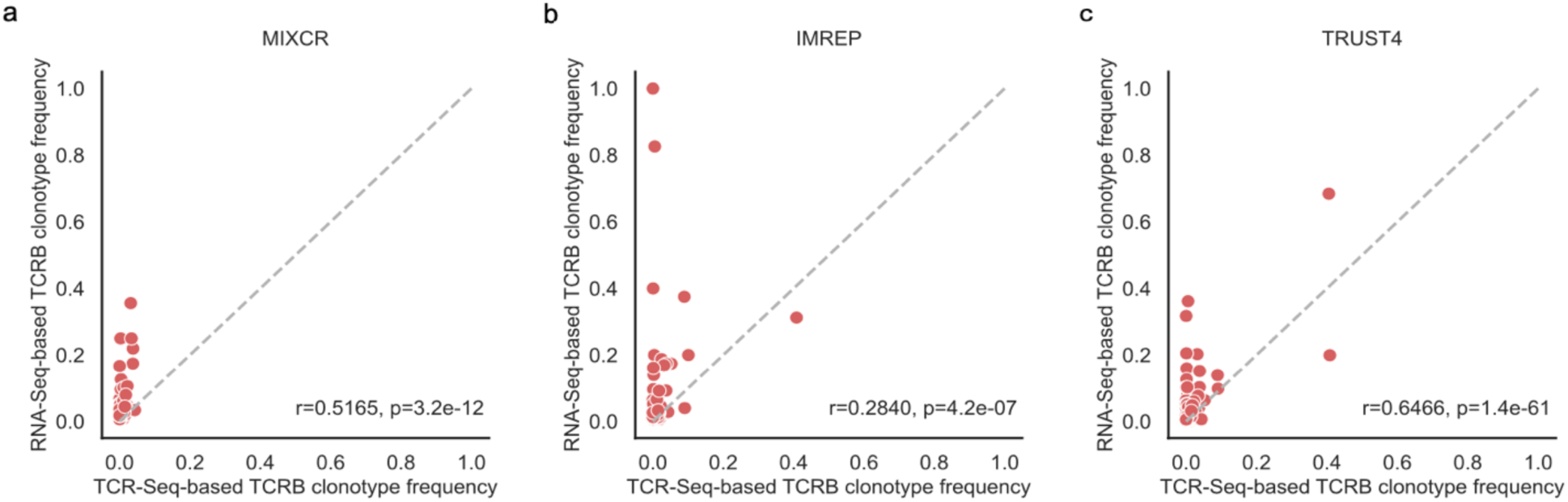
Correlation of TCRβ clonotype frequency based on the TCR-Seq data (x-axis) and the TCR derived reads from RNA-seq (y-axis) in T cell poor polyclonal samples. Only clonotypes assembled from RNA-Seq data are presented. Pearson correlation coefficients and the corresponding p-values are calculated and reported.

**Supplementary Fig.12.**
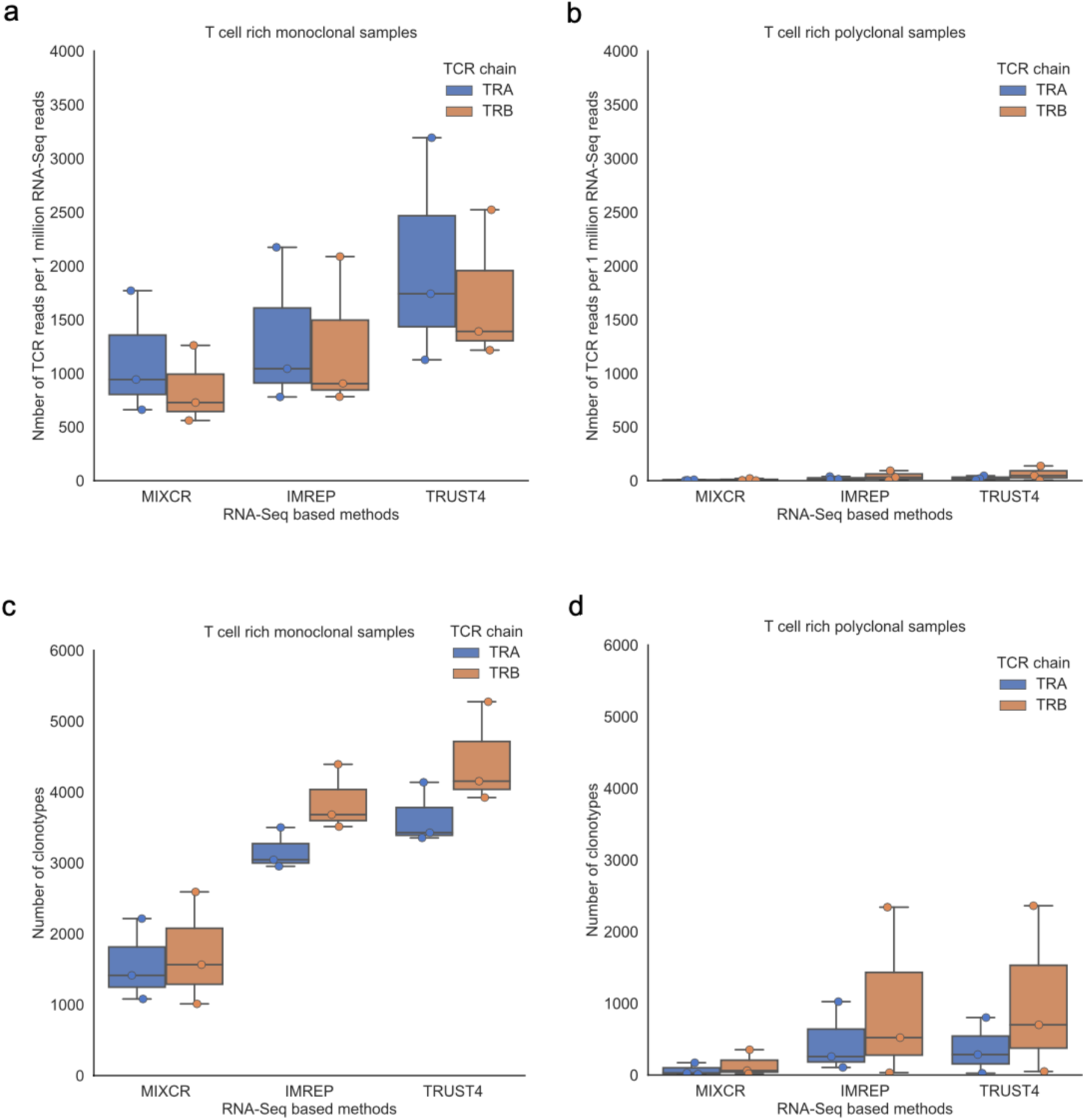
TCRα chain and TCRβ chain metrics comparison. a-b. TCR derived reads from RNA-Seq reads per one million RNA-Seq reads in T cell rich monoclonal samples (a) and T cell rich polyclonal samples (b). c-d. TCRα and TCRβ chain clonotype counts in T cell rich monoclonal samples (c) and T cell rich polyclonal samples (d). The results from TCRα chain are shown in blue and the results from TCRβ are shown in orange.

